# Quantitative bias analysis in practice: Review of software for regression with unmeasured confounding

**DOI:** 10.1101/2022.02.15.22270975

**Authors:** E Kawabata, K Tilling, RHH Groenwold, RA Hughes

**Affiliations:** MRC Integrative Epidemiology Unit, University of Bristol, Bristol, United Kingdom; Population Health Sciences, Bristol Medical School, University of Bristol, Bristol, United Kingdom; Department of Clinical Epidemiology, Leiden University Medical Center, Leiden, The Netherlands; Department of Biomedical Data Sciences, Leiden University Medical Center, Leiden, The Netherlands

**Keywords:** Causal inference, Linear regression, Review, Sensitivity analysis, Software, Unmeasured confounding

## Abstract

Failure to appropriately account for unmeasured confounding may lead to erroneous conclusions. Quantitative bias analysis (QBA) can be used to quantify the potential impact of unmeasured confounding or how much unmeasured confounding would be needed to change a study’s conclusions. Currently, QBA methods are not routinely implemented, partly due to a lack of knowledge about accessible software. We review the latest developments in QBA software between 2011 to 2021 and compare five different programs applicable when fitting a linear regression: *treatSens, causalsens, sensemakr, EValue*, and *konfound*. We illustrate application of these programs to two datasets and provide code to assist analysts in future use of these software programs. Our review found 21 programs with most created post 2016. All are implementations of a deterministic QBA, and the majority are available in the free statistical software environment R. Many programs include features such as benchmarking and graphical displays of the QBA results to aid interpretation. Out of the five programs we compared, *sensemakr* performs the most detailed QBA and includes a benchmarking feature for multiple unmeasured confounders. The diversity of QBA methods presents challenges to the widespread uptake of QBA among applied researchers. Provision of detailed QBA guidelines would be beneficial.

## 1 Introduction

The main aim of many epidemiology studies is to estimate the causal effect of an exposure on an outcome (here onward, shortened to exposure effect). In observational studies participants are not randomised to exposure (or treatment) groups. Consequently, factors that affect the outcome are typically unevenly distributed among the exposure groups, and a direct comparison between the exposure groups will likely be biased due to confounding. Standard adjustment methods (such as standardization, inverse probability weighting, regression adjustment, g-estimation, stratification and matching) assume the adjustment model is correct and a sufficient set of confounders has been measured without error^1^. Failure to appropriately account for unmeasured or poorly measured confounders in analyses may lead to invalid inference^2–4^.

There are several approaches to assess causality which depend on assumptions other than “no unmeasured confounding” (e.g., self-controlled study designs, prior event rate ratio, instrumental variable analysis, negative controls, perturbation variable analysis, and methods that use confounder data collected on a study sub-sample^5^). When none of these approaches are applicable (e.g., study lacks an appropriate instrument or sub-sample data on the unmeasured confounders) then the analyst must assess the sensitivity of the study’s conclusions to the assumption of no unmeasured confounding using a quantitative bias analysis (QBA; also known as a sensitivity analysis). A QBA can be used to quantify the potential impact of unmeasured confounding on an exposure effect estimate or to quantify how much unmeasured confounding would be needed to change a study’s conclusions.

Currently, QBA methods are not routinely implemented. A recent published in 2016 found that the use of QBA for unmeasured confounding had not increased in the years 2010 − 2012 compared to the 2004 − 2007 period^6^. Lack of knowledge about QBA, and of analyst-friendly methods and software have been identified as barriers to the widespread implementation of a QBA^7–9^. In the past decade, there have been several reviews of QBA methods^2,5,9–17^. Only two of these papers reviewed software implementations of QBA methods: the supplementary of^15^ provided a brief summary of software implementing Rosenbaum-style QBA methods^18^, and^11^ reviewed software implementations before its publication in July 2014. Also, comparisons of QBA methods have primarily been limited to analyses with a binary outcome^10,19–26^.

Our paper reviews available software implementing a QBA to address unmeasured confounding caused by a study not collecting data on these confounders as opposed to mismeasurement of measured confounders. We then describe, illustrate and compare QBA software applicable when the analysis of interest is a linear regression. We illustrate how to apply these methods using a real-data example from the Barry Caerphilly Growth (BCG) study^27,28^, and, in the Supplementary Materials, we also provide code implementing these methods when applied to publicly-accessible data from the 2015 − 2016 National Health and Nutrition Examination Survey (NHANES) study^29^.

## 2 Quantitative bias analysis for unmeasured confounding

We want to estimate the effect of an exposure (or treatment) *X* on an outcome *Y*. The *Y* − *X* association is confounded by measured covariates *C* and unmeasured confounders *U*. The naive estimate of the exposure effect, 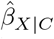, assumes no unmeasured confounding and is estimated by controlling for *C* only.

We can use a QBA to quantify the likely magnitude and direction of the bias, due to unmeasured confounding, under different plausible assumptions about *U* (assuming no other sources of bias). Generally, a QBA requires a model (known as a bias model) for the observed data, *Y, X* and *C*, and unmeasured data, *U*. The bias model will include one or more parameters (known as bias or sensitivity parameters) which cannot be estimated from the observed data. Therefore, values for these bias parameters must be prespecified before conducting the QBA. Typically, the bias parameters specify the strength of the association between *U* and *X* given *C*, and between *U* and *Y* given *X* and *C*^21^. Information about the likely values of these bias parameters may be obtained from external sources (such as external validation studies, published literature, or expert opinion)^8^, and from benchmarking (also known as calibration) where strengths of associations of measured covariates *C* with *X* and *Y* are used as benchmarks^30^ for the bias parameters. We shall denote the bias parameters by *ϕ* and the bias-adjusted estimate of the exposure effect assuming *ϕ* by 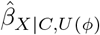.

A QBA is often conducted as a tipping point analysis, where the analyst identifies the values of *ϕ* that correspond to a change in the study conclusions (known as the “tipping point”). A tipping point analysis may be applied to the point estimate or confidence interval (CI) of the exposure effect; for example, to identify the values of *ϕ* corresponding to a null effect, or the values of *ϕ* corresponding to a statistically insignificant effect of a non-null point estimate. If the values of *ϕ* at the tipping point(s) are considered unlikely then the study conclusions are said to be robust to unmeasured confounding.

There are two broad classes of QBA methods: deterministic and probabilistic^7^. A deterministic QBA specifies a range of values for each bias parameter of *ϕ* and then calculates 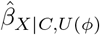 for all combinations of the prespecified values of *ϕ*. Typically, the results are displayed as a plot or table of 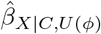 against different values of *ϕ*. Unlike a deterministic QBA, a probabilistic QBA uses a prior probability distribution for *ϕ* to explicitly model the analyst’s assumptions about which combinations of *ϕ* are most likely to occur and to incorporate their uncertainty about *ϕ*^7,22^. Averaging over this probability distribution generates a distribution of estimates of 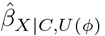 which is summarised to give a point estimate (i.e., the most likely 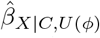 under the QBA’s assumptions) and an interval estimate (i.e., defined to contain the true exposure effect with a prespecified probability) which accounts for uncertainty due to the unmeasured confounding and sampling variability^7^.

## 3 Overview of available software

The aim of the literature review was to give a brief overview of publicly available software implementations of QBA, described in articles published between 1st January 2011 and 31st December 2021 (inclusive). We defined ”software” to be either a web tool with a user-interface or software code that (i) was not specific to a particular data example (i.e., we excluded examples of code from empirical analyses), (ii) was freely available to download, and (iii) was accompanied by documentation detailing the software’s features.

Our literature search was conducted in three stages. In stage 1, we used Web of Science to identify papers that mentioned “quantitative bias analysis” and “unmeasured confounding” (or their synonyms) in either the title, abstract or as keywords (see Supplementary Box 1 for our search strategy). In stage 2, the abstracts were reviewed by two independent reviewers to determine if they were eligible for data extraction with any disagreements resolved by consensus. Eligible abstracts were published articles that either introduced a new QBA method or software implementation, compared or reviewed existing QBA methodology, or gave a tutorial on QBA. Examples of ineligible abstracts were meeting abstracts, commentaries, articles where a QBA was not conducted but mentioned as further work, and articles solely focused on answering applied questions (and so included limited information on the statistical methodology used). In stage 3, we extracted from the full text information about the analysis of interest, the QBA method, and the software used to implement the QBA.

After excluding duplicates, our Web of Science search identified 780 papers (flowchart of the review shown in Supplementary Figure S1). We excluded 24 meeting abstracts and editorials, 379 articles that did not conduct a QBA to unmeasured confounding, and 239 articles on applied analyses. Of the remaining 138, 29 articles referred to 21 publicly available software implementations of a QBA.

Table 1 summarises the key features of the 21 software programs in ascending date-order of creation. All 21 programs implement a deterministic QBA, with only 8 programs publicly available before 2017, and 17 implemented in the free software environment R^31^. Seven programs implement a QBA applicable for a matched observational study, five for a mediation analysis, and nine for a standard regression analysis. Five of the seven programs for a matched analysis (*sensitivityCaseControl, sensitivitymw, sensitivitymv, sensitivityfull* and *submax*) implement the same QBA method^18,32^ but for different types of matched observational studies. For example, *sensitivitymw* is applicable to matched sets with one exposed subject and a fixed number of unexposed subjects, and *sensitivitymv* to matched sets with one exposed subject and a variable number of unexposed subjects. Also, *submax* and *sensitivityCaseControl* exploit effect modification and different definitions of a case of disease, respectively, to further evaluate sensitivity to unmeasured confounding. Among the programs for mediation analysis, *MediationSensitivityAnalysis* evaluates sensitivity to unmeasured confounding of the mediator-outcome relationship only, while the remaining programs can also evaluate sensitivity to unmeasured confounding of the exposure-mediator and exposure-outcome relationships.

**Table 1.**
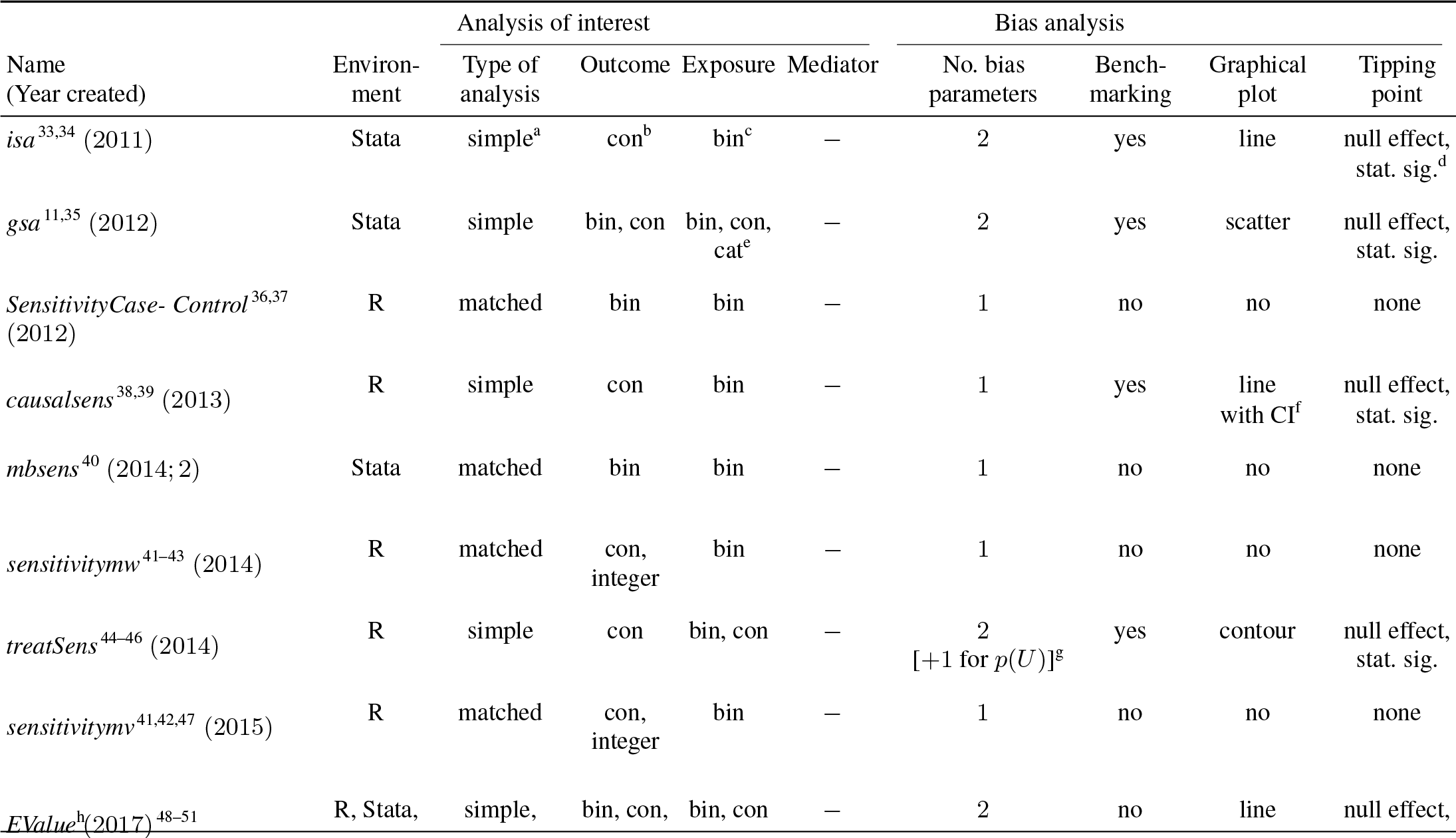
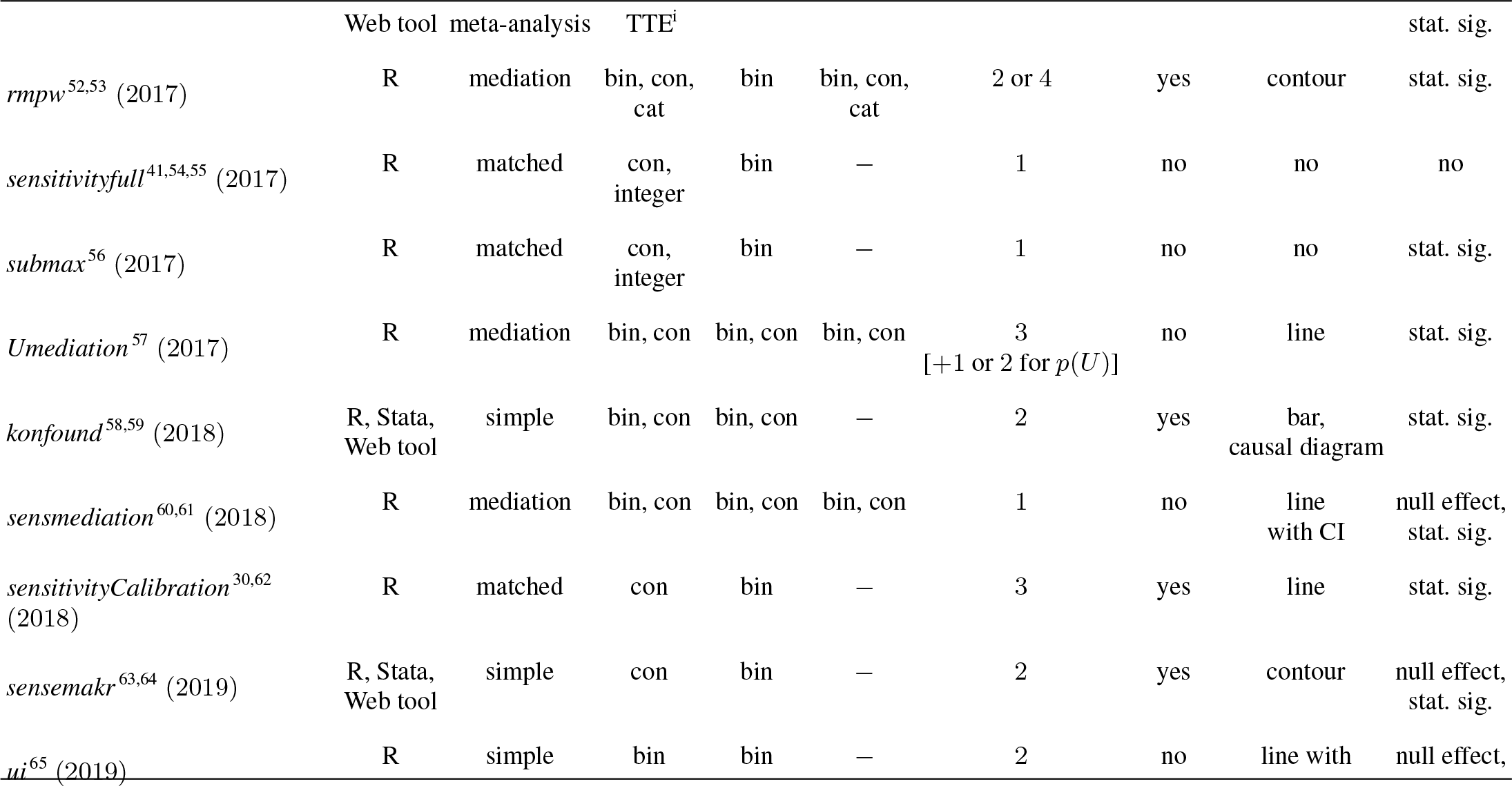
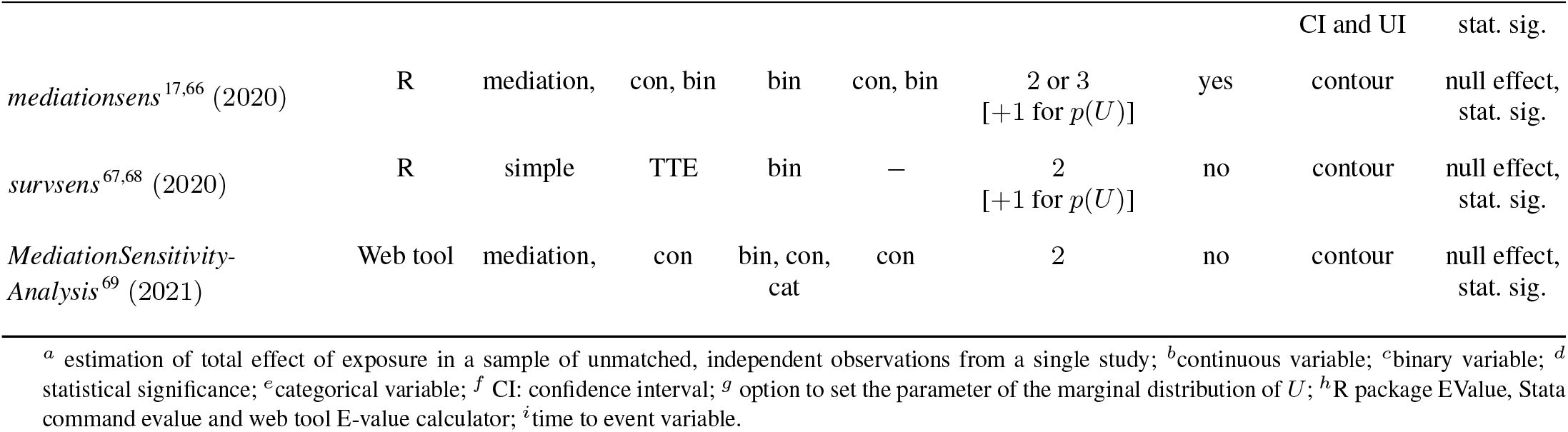
Software programs implementing a quantitative bias analysis for unmeasured confounding reported in articles published between 2011 and 2021.

Most programs require the outcome (of the analysis of interest) to be either binary or continuous. However, program *survsens* implements a QBA specifically for a Cox proportional hazards regression analysis and is applicable for survival outcomes with or without competing risks. All programs can be applied to a binary exposure and seven programs are also applicable to a continuous or categorical exposure. Also, all programs allow the analysis of interest to adjust for measured covariates *C* of any variable type and generally assume that *U* represents the part of the unmeasured confounder(s) that is independent of *C*. Nine programs use the measured covariates to calculate benchmark values for the bias parameters.

The bias parameters represent the strength of the relationships between *U* and the exposure, outcome, or mediator. Programs *treatSens, Umediation, mediationsens*, and *survsens* also allow the analyst to vary the parameters of the marginal distribution of *U* (e.g., for binary *U* the probability Pr(*U* = 1)). Otherwise, these marginal parameters are set to a default value (e.g., Pr(*U* = 1) = 0.5).

Almost all programs report the values of the bias parameters at prespecified tipping points. Also, most programs output the bias-adjusted results (e.g., point estimate, CI or P-value for the exposure effect) at prespecified values of the bias parameter(s) (exceptions include *isa, gsa, konfound*, and R and Stata implementations of *EValue*). Note that, programs *uMediation* and *ui* summarise the bias-adjusted results using uncertainty intervals, which incorporates uncertainty about the values of the bias parameters and sampling variability. Fifteen programs generate a graphical plot of their QBA results.

Two programs also implement a QBA to other sources of bias: *MediationSensitivty-Analysis* can assess sensitivity to measurement error of the mediatior, outcome and measured covariates, and *Evalue* can assess sensitivity to differential misclassification of an outcome or exposure and to sample selection bias. Furthermore, both programs can simultaneously assess sensitivity to multiple sources of bias.

## 4. Quantitative bias analysis methods for linear regression

We describe and illustrate the following programs from Table 1 applicable for an unmatched analysis, where the exposure is binary and the exposure effect is estimated by a linear regression model: *treatSens*^44,45^, *causalsens*^38^, *sensemakr*^70^, *EValue*^48^, and *konfound*^58^. For reasons of brevity, we excluded programs *isa* and *gsa* as they are similar to the more recently published *treatSens*.

All five programs are implemented as an R package^39,46,49,59,64,71,72^, and *sensemakr*^64^, *EValue*^49,50^ and *konfound*^58,73^ are also available as a Stata command and web tool. Individual participant data is required for *treatSens* and *causalsens, EValue* only requires summary data from the naive analysis, and *sensemakr* and *konfound* can be applied to individual participant and summary data. Additionally, *treatSens, causalsens*, and *sensemakr* require prespecified values for *ϕ* which can be set by the analyst or set using the program’s default values. Note that all five methods can be applied when 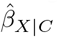 is not null, irrespective of whether 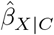 is statistically significant or not, and when 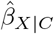 is null. However, for *treatSens* the tipping point for the point estimate is fixed at the null, and so this feature can only be used when 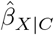 is not null. Below is a summary of the five programs with further details in the Supplementary Materials.

### 4.1 treatSens

Program *treatSens* implements a simulation-based QBA^44^ which is similar to multiple imputation for missing data^74^. For a prespecified value of *ϕ, treatSens* simulates *U* multiple times from the conditional distribution *U Y, X, C* given by the bias model. For each set of simulated values of *U*, the exposure effect is estimated from a linear regression of *Y* given *X, C* and the simulated *U*, and then Rubin’s rules^74^ are used to combine the multiple sets of results into a single estimate for 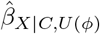 and its standard error.

The bias model consists of three sub-models: the analysis model (i.e., linear regression of *Y* given *X, C* and *U*), the treatment model (e.g., linear or probit regression regression of *X* on *C* and *U* for continuous or binary *X*, respectively), and a marginal model for *U* (standard normal or Bernoulli distribution for continuous or binary *U*, respectively). It has two bias parameters *ϕ* = (*ζ*^*Y*^, *ζ*^*Z*^): *ζ*^*Y*^ is the coefficient for *U* from the analysis model *Y X, C, U* and *ζ*^*Z*^ is the coefficient of *U* from the treatment model *X C, U*. To allow for bias in both directions (i.e., increased exposure effect, and reduced or reversed exposure effect), positive and negative values are specified for *ζ*^*Z*^. The remaining coefficients of the treatment and analysis models are estimated from the observed data. The coefficients of measured covariates *C* (from the regressions of *Y* on *X* and *C*, and *X* on *C*) are used as benchmark values for *ζ*^*Y*^ and *ζ*^*Z*^, respectively^44^. All continuous variables are standardised to facilitate comparison between these benchmark values and the bias parameters.

Program *treatSens* outputs a contour plot of the bias-adjusted estimates, 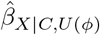, for different combinations of *ζ*^*Y*^ and *ζ*^*Z*^, indicating the values of *ζ*^*Y*^ and *ζ*^*Z*^ that correspond to tipping points for the point estimate (fixed at the null) and statistical significance (analyst can set the significance level; default is 5%). Additional outputs include tables of: (1) combination values of *ζ*^*Y*^ and *ζ*^*Z*^ at the tipping points, (2) 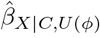 and corresponding standard errors for prespecified values of *ζ*^*Y*^ and *ζ*^*Z*^ and each set of simulated *U*, and (3) benchmark values for *ζ*^*Y*^ and *ζ*^*Z*^.

### 4.2 causalsens

Program *causalsens* generates a modified outcome, 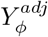, which is adjusted for the bias due to unmeasured confounding for a prespecified value of *ϕ*^38^. The naive analysis is then refitted using 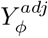 instead of *Y* and the resulting exposure effect estimate and CI are the bias-adjusted results.

The QBA of *causalsens* is based on the potential outcomes framework^75^. Program *causalsens* requires a binary *X*, and so there are two potential outcomes per subject: *Y* (0) when not exposed and *Y* (1) when exposed. The bias model consists of a treatment model and a “confounding function”^76,77^. The treatment model is a logistic regression used to estimate the probability of being in the exposed group given *C*. The confounding function quantifies the average difference in potential outcomes *Y* (0) (or *Y* (1)) between those in the exposed and unexposed groups, with any nonzero difference attributed to unmeasured confounding. It is parameterised by a single bias parameter, 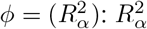 denotes the proportion of unexplained variance in the potential outcomes that is explained by *U* and is set to positive and negative values to allow *U* to move the point estimate towards and away from the null. Program *causalsens* supplies two choices for the confounding function, named the “one-sided function” and the “alignment function”, and also allows the analyst to specify their own function. The one-sided function assumes the true exposure effect is identical in the exposed and unexposed groups. Setting 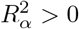 implies that the mean of *Y* (1) (and *Y* (0)) is higher for the exposed group than the unexposed group, leading 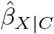 to be positively biased; and vice versa for 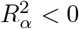 (See the Supplementary Materials for details of the alignment function.)

Program *causalsens* outputs a line plot and a table of 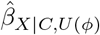 and corresponding 95% CI for different values of 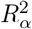. Additionally, *causalsens* reports benchmarks for 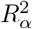 based on the partial *R*^2^ values for each covariate in *C*.

### 4.3 sensemakr

Program *sensemakr* uses formulae to estimate 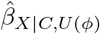 and its t-value for prespecified values of *ϕ*. Additionally, *sensemakr* reports summary measures, called “robustness values”, which quantify the minimum amount of unmeasured confounding needed to change a study’s conclusions, conditional on *C*^63^.

The bias model of *sensemakr* expresses the absolute difference between the naïve and bias-adjusted estimates, 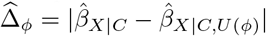, and the standard error of 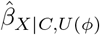 as functions of estimated quantities from the naive analysis and 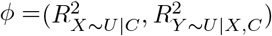. Bias parameter 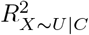 is the proportion of the variance of *X*, not explained by *C*, that is explained by *U*, and 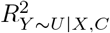 is the proportion of the variance of *Y*, not explained by *X* and *C*, that is explained by *U*. Considering both directions of effect of *U*, 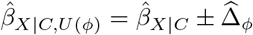, and the corresponding t-value for a null hypothesis is 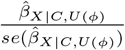.

The robustness value for the point estimate (or t-value) represents the minimum value of 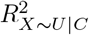 and 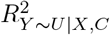, when 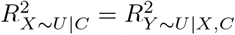, such that 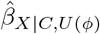 (or its t-value) equals its prespecified tipping point value; for example, the null (or the 5% critical t-value). A robustness value close to 1 indicates that strong unmeasured confounding would be needed to change the study conclusions, whilst a value close to 0 indicates that very weak unmeasured confounding could change the conclusions.

Program *sensemakr* calculates upper bounds (called “benchmark bounds”) for 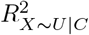 and 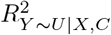 using *C*^63,70^. The benchmark bounds based on covariate *C*_*j*_ represent the maximum values for 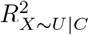 and 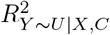 if *U* was *k* times (*k* = 1, 2, 3, …) as strong as *C*_*j*_ (in terms of strengths of relationships with *X* and *Y*)^63^. Additionally, *sensemakr* can calculate benchmark bounds based on a group of measured covariates.

Program *sensemakr* outputs robustness values for the point estimate and t-value, and contour plots of 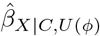 and corresponding t-value for prespecified values of *ϕ*, indicating the combinations of 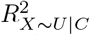 and 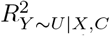 that correspond to a tipping point for the point estimate or t-value. Also, *sensemakr* outputs a table of benchmark bounds and values of 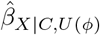 and corresponding CI when 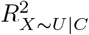 and 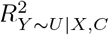 equals these benchmark bounds. Note that, only the R package and Stata command can calculate benchmark bounds based on more than one measured covariate.

### 4.4 EValue

Program *EValue* reports a summary measure, called an E-value, which quantifies the minimum amount of unmeasured confounding needed to change a study’s conclusions, conditional on the measured covariates^48^. The E-value is defined on the risk ratio scale and is a function of estimated quantities from the naive analysis and two bias parameters *ϕ* = (*RR*_*XU*_, *RR*_*UY*_). For binary *X* and a single, binary *U, RR*_*XU*_ represents the risk ratio for the effect of *X* on *U* conditional on *C* and *RR*_*UY*_ represents the maximum risk ratio for the effect of *U* on *Y* after adjustment for *C* among the exposed and unexposed^78^. (See Ding and VanderWeele^78^ for a definition of *RR*_*XU*_ and *RR*_*UY*_ when *U* denotes a single or multiple unmeasured confounders of type continuous, categorical or mixed.) For effect measures other than the risk ratio, the naive results are first converted to the risk ratio scale before calculating the E-value^48^. For example, for standardised mean difference, 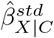, and corresponding standard error, 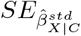, the approximate risk ratio for the point estimate is 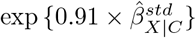 and the approximate risk ratio for a limit of the 95% CI is 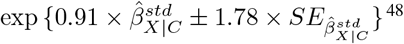^48^. Note that, the E-value is interpreted on the risk ratio scale for all types of effect measures.

Here we describe the E-value when the tipping point of the point estimate is the null, although it can also be set to a non-null value (see Supplementary Materials of^48^). A separate E-value is calculated for the point estimate and CI limit closest to the null. The E-value for the point estimate (or CI limit) represents the minimum value of *RR*_*XU*_ and *RR*_*UY*_, when *RR*_*XU*_ = *RR*_*UY*_, such that 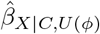 is null or in the reverse direction to that of 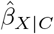 (or the exposure effect is no longer statistically significant after adjustment for *C* and *U*). The E-value is a positive number *≥* 1 with higher values indicating that greater levels of unmeasured confounding (i.e., stronger *X* − *U* and *Y* − *U* associations) are required to change the study conclusions. When 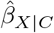 is null (or its CI includes the null) the E-value for the point estimate (or CI limit) is 1, indicating that no unmeasured confounding is required to change the study conclusions. Importantly, the E-value is a measure of sensitivity to unmeasured confounding for a worst-case scenario (i.e., bias parameters *RR*_*XU*_ and *RR*_*UY*_ are set to values which maximize the bias due to unmeasured confounding)^79^.

The R package *EValue*, Stata command *evalue*, and web tool *e-value calculator* can all be applied when the effect measure of interest is a risk ratio, risk difference, standardised mean difference, odds ratio or hazard ratio for a rare outcome (i.e., prevalence *<* 15%), and odds ratio or hazard ratio for a common outcome (i.e., prevalence *≥* 15%). From here onward, we shall use *EValue* to represent all three implementations. Program *EValue* outputs E-values for the point estimate and CI limit, and a line plot of the values of *RR*_*UY*_ and *RR*_*XU*_ that correspond to prespecified tipping points for the point estimate and CI limit. Note that, the program does not supply benchmark values for *RR*_*UY*_ and *RR*_*XU*_. For comparison purposes, VanderWeele and Ding^48^ suggest omitting each measured covariate in turn and recalculating the E-value.

### 4.5 konfound

Program *konfound* assesses sensitivity to a change in the statistical (in)significance status of 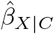^58^. This includes the scenario where *U* explains away all of the statistical significance of 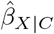 (i.e., 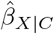 is statistically significant but 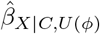 is statistically insignificant) and the converse scenario where *U* restores the statistical significance of 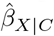 (i.e., 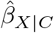 is statistically insignificant but 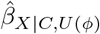 is statistically significant). Program *konfound* refers to the first scenario as *U* “invalidating inference” and the second as *U* “sustaining inference”. By default, the significance level is 5% and the null hypothesis is “no exposure effect”, both of which can be changed by the analyst.

Program *konfound* reports two summary measures that quantify the minimum level of unmeasured confounding necessary to change conclusions on statistical significance: percent bias and impact threshold. Percent bias is a measure of the minimum percentage 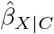 of that would need to be explained away by *U* in order for unmeasured confounding to invalidate inference^80,81^. The formula for the percent bias is a function of estimated quantities from the naive analysis and the value of 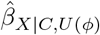 when its P-value is exactly *κ*% (for statistical significance defined at the *κ*% level). The impact threshold is also derived from estimated quantities of the naive analysis plus two bias parameters *ϕ* = (*r*_*X*∼*U*|*C*_, *r*_*Y* ∼*U*|*C*_): *r*_*X*∼*U*|*C*_ and *r*_*Y* ∼*U*|*C*_ represent the partial correlation between *U* and *X* and between *U* and *Y* (conditional on *C*), respectively^82^. The impact threshold is the product *r*_*X*∼*U*|*C*_ × *r*_*Y* ∼*U*|*C*_ when *r*_*X*∼*U*|*C*_ and *r*_*Y* ∼*U*|*C*_ are equal and set to their minimum value such that statistical inference is invalidated or sustained. Note that, the percent bias measure is always positive but the impact threshold measure can be positive or negative depending on the direction of the correlation between *U* and *X* and *Y*. For both measures, larger absolute values indicate greater robustness to unmeasured confounding. Program *konfound* calculates the impact threshold and percent bias when *Y* is a continuous outcome and the naive analysis is a linear regression.

The software outputs the percent bias (depicted by a bar graph called a “threshold plot”) and the impact threshold (depicted by a causal-type diagram called a “correlation plot”; generated by the R package and online tool). Only the Stata command provides benchmark values for *r*_*X*∼*U*|*C*_ and *r*_*Y* ∼*U*|*C*_, which are the partial correlation of each measured covariate *C*_*j*_ with *X* and with *Y*, respectively, given the remaining measured covariates.

## 5 Illustrative example

We applied the five QBA methods of Section 4 to data from the BCG and NHANES studies. For both examples, the naive analysis was the linear regression *Y* | *X, C* with binary exposure *X*. We used measured variables to represent the unmeasured confounders *U*. So, in effect our analyses examined the effect of not including certain confounders and we assumed that after adjustment for *U* and *C* there was no unmeasured confounding. In the BCG example, *U* was a single confounder and adjustment for *U* did not change the study conclusions. See the Supplementary Materials for the NHANES example where *U* represents multiple confounders.

For *treatSens* we used Probit regression for its treatment model because *X* was binary, and for *causalsens* we used the one-sided confounding function because we assumed the exposure effect was the same in both exposure groups.

Using *C*, we calculated benchmark E-values and, for the other four programs, we calculated benchmark values for *ϕ* and the bias-adjusted results when *ϕ* was set to (multiples of) the benchmark values corresponding to the “strongest measured covariate” (i.e., the covariate that had the strongest associations with *X* and *Y*).

As this is an illustrative example of applying a QBA to unmeasured confounding, we have ignored other potential sources of bias (such as missing data) and only considered a small number of measured covariates. We restricted our analyses to participants with complete data on *Y, X, C* and *U*.

### 5.1 Description of the BCG Study

The BCG study is a follow-up of a dietary intervention randomized controlled trial of pregnant women and their offspring^27,28^. Data were collected on the offspring (gestational age, sex, and 14 weight and height measures at birth, 6 weeks, 3, 6, 9 and 12 months, and thereafter at 6-monthly intervals until aged 5 years) and their parents (anthropometric measures, health behaviours and socioeconomic characteristics). When aged 25, these offspring were invited to participate in a follow-up study in which standard anthropometric measures were recorded. We refer to the offspring, later young adults in the follow-up study, as the study participants.

Our analysis was a linear regression of adult body mass index (BMI) at age 25 on being overweight at age 5 years (BMI *≥* 17.44 kg/m^2 83^). Measured covariates *C* were participant’s sex, gestational age, birth weight, and parents’ height and weight measurements. The strongest measured covariate was maternal weight. The unmeasured confounder *U* was a measure of childhood socioeconomic position (SEP) (paternal occupational social class based on the UK registrar general classification^84^) with *U* = 1 for professional or managerial occupations, and *U* = 0 otherwise. Based on the 542 participants with complete data on all variables, 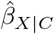 was 2.21 kg/m^2^ (95% CI 1.30, 3.11 kg/m) and the fully adjusted estimate (i.e., adjusted for *C* and *U*) was 2.19 kg/m^2^ (95% CI 1.29, 3.09 kg/m^2^). Also, the coefficient of *U* from the linear regression *Y* | *X, C, U* was − 0.66 kg/m^2^ (95% CI − 1.57, 0.25 kg/m^2^) and the coefficient of *U* from the logistic regression *X* | *C, U* was − 0.23 (95% CI − 0.85, 0.35). Statistical significance was defined at the 5% level.

Note that, on a computer with 2.7 Ghz the run-time of *treatSens* (with the default setting of single-threading^46^) was 10 minutes while the other programs generated their results instantaneously. We begin with a description of the outputted results and then compare the results across the five programs. In the Supplementary Materials we report on a small survey we conducted to obtain feedback on how the five QBA programs compare with respect to ease/difficultly of interpreting their QBA results.

### 5.2 Results

#### treatSens

Program *treatSens* outputs a contour plot (Figure 1(a)) where each contour represents the different combinations of *ϕ* = (*ζ*^*Y*^, *ζ*^*Z*^) that result in the same bias-adjusted estimate, 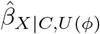. For example, 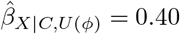 standard deviations of BMI (SD-BMI; or equivalently 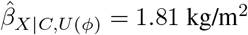) when *ζ*^*Y*^ = 0.23 and *ζ*^*Z*^ = 1.00, and when *ζ*^*Y*^ = 1.00 and *ζ*^*Z*^ = 0.24. (Note that, *treatSens* standardises all continuous variables.) The black horizontal contour at *ζ*^*Y*^ = 0 denotes the naive estimate of 0.49 SD-BMI (i.e., 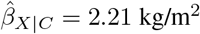), the red contour represents the combinations of *ϕ* that would result in a null exposure estimate, and the blue contours bracket statistically insignificant exposure estimates. The pluses and inverted triangles denote the benchmark values of *ϕ* based on measured covariates *C*: pluses represent covariates positively associated with adult BMI, and the inverted triangles represent covariates negatively associated with adult BMI with those negative associations rescaled by − 1. The red cross furthest away from the origin denotes the strongest measured covariate (maternal weight).

**Figure 1.**
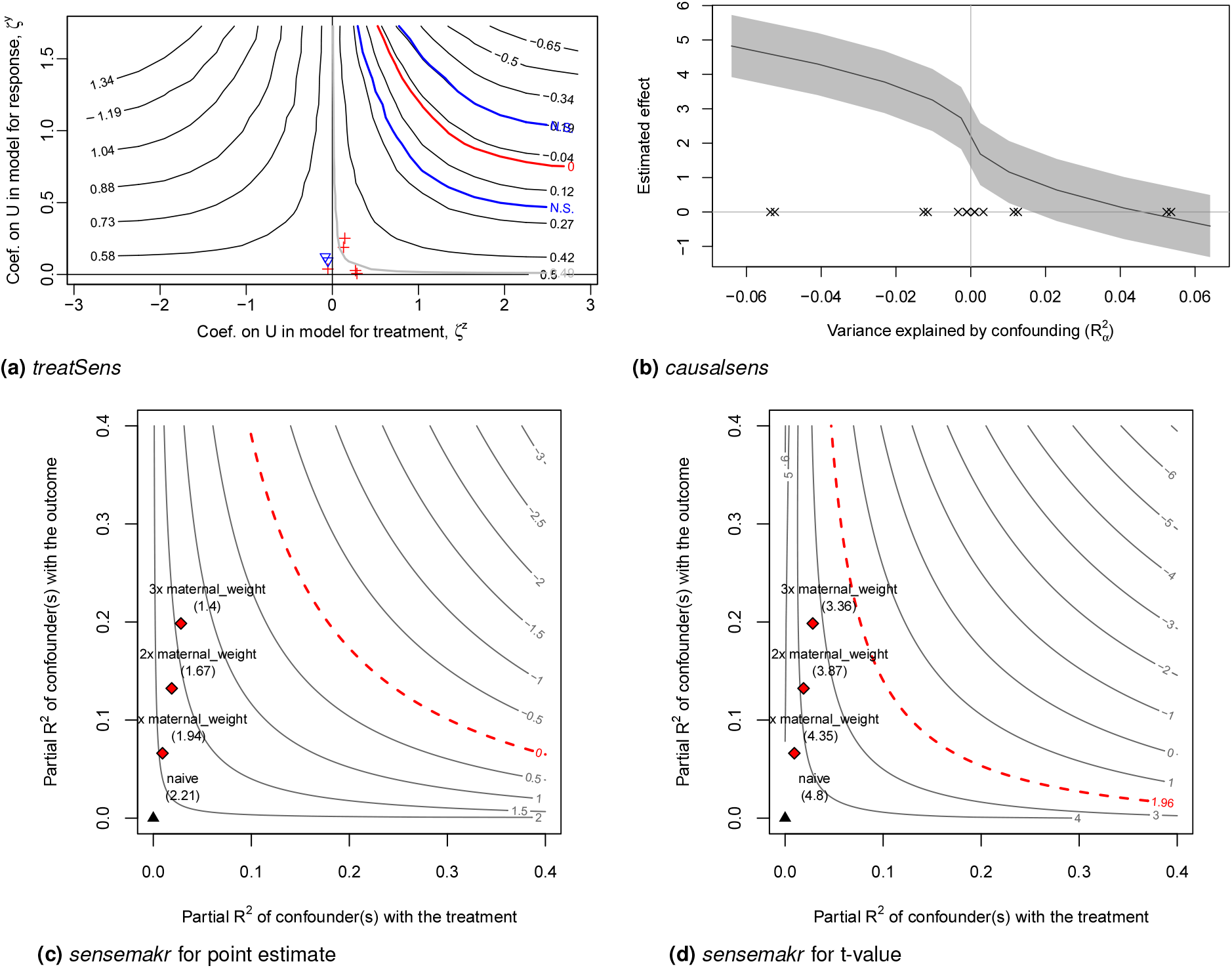
Quantitative bias analysis for effect of child overweight on adult body mass index from the Barry Caerphilly Growth study. Red contour (null effect in (a) and (c), t-value at 5% significance in (d)), blue contours (bracket 5% statistically insignificant estimates), black contour or line (bias-adjusted estimates), grey shaded area (95% confidence intervals for bias-adjusted estimates), pluses, inverted triangles, crosses, and diamonds (benchmarks), and black triangle (naive estimate).

#### causalsens

Program *causalsens* outputs a line plot (Figure 1(b)) where the black line represents the bias-adjusted exposure estimates, the grey shaded area represents the corresponding 95% CIs, and the crosses denote the benchmark values for 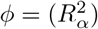 with each benchmark appearing twice to allow for both directions of effect. Values of 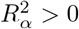 implies that individuals in the unexposed group tended to be healthier (i.e., lower adult BMI) than those in the exposed group even if everyone was of normal weight (or overweight) at age 5; and the converse for 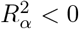.

#### sensemakr

Program *sensemakr* outputs four contour plots: Figures 1(c) and (d) show the contour plots for the exposure effect estimate and its t-value, respectively, generated under the assumption that accounting for *U* moves the exposure effect estimate closer to the null, and Supplementary Figures S2(a) and (b) show the same contour plots generated under the converse assumption. The contours have a similar interpretation as discussed for *treatSens*. For example, the red contour represents different combinations of 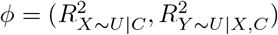 that result in a null exposure effect (Figure 1(c)) and the critical t-value corresponding to 5% statistical significance (Figure 1(d)). The black triangle denotes the naive estimate, 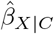, and the red diamonds denote once, twice and thrice the benchmark bounds based on the strongest measured covariate.

The robustness values for 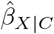 and its t-value were 18.76% and 11.56%, respectively. Thus, *U* would need to explain at least 18.76% (or 11.56%) of the residual variance of both childhood overweight and adult BMI for the exposure effect adjusted for *C* and *U* to be null (or statistically insignificant).

#### EValue

The E-values for 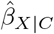 and its lower CI limit were 2.50 and 1.93, respectively. Thus, if the associations between *U* and adult BMI and childhood overweight were at least 2.50 (or 1.93), on the risk ratio scale, then the exposure effect adjusted for *C* and *U* may be null or in the reverse direction (or strictly positive but statistically insignificant). Supplementary Figure S3 shows the combinations of *ϕ* = (*RR*_*UY*_, *RR*_*XU*_) that correspond to a null bias-adjusted estimate (red contour) and a strictly positive but statistically insignificant bias-adjusted estimate (black contour).

#### konfound

The percent bias was 59.11%, depicted in the bar-graph shown in Supplementary Figure S4, and the impact threshold was 0.13 with bias parameters 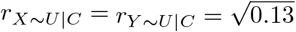, depicted in the causal diagram shown in Supplementary Figure S5. Therefore, in order for the exposure effect to be statistically insignificant after adjustment for *C* and *U* then (1) *U* would need to account for at least 59.11% of 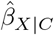 (i.e., 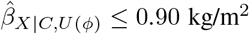), and (2) the partial correlations of *U* with adult BMI and child overweight must both exceed 0.36.

#### Comparison of the results

Table 2 summarises the bias-adjusted results of each program in scenarios where the associations between *U* and adult BMI and child overweight were half, once and twice as strong as the corresponding associations with the strongest measured covariate (i.e., *ϕ* set to 0.5, 1 and 2 *×* benchmark values for maternal weight).

**Table 2.**
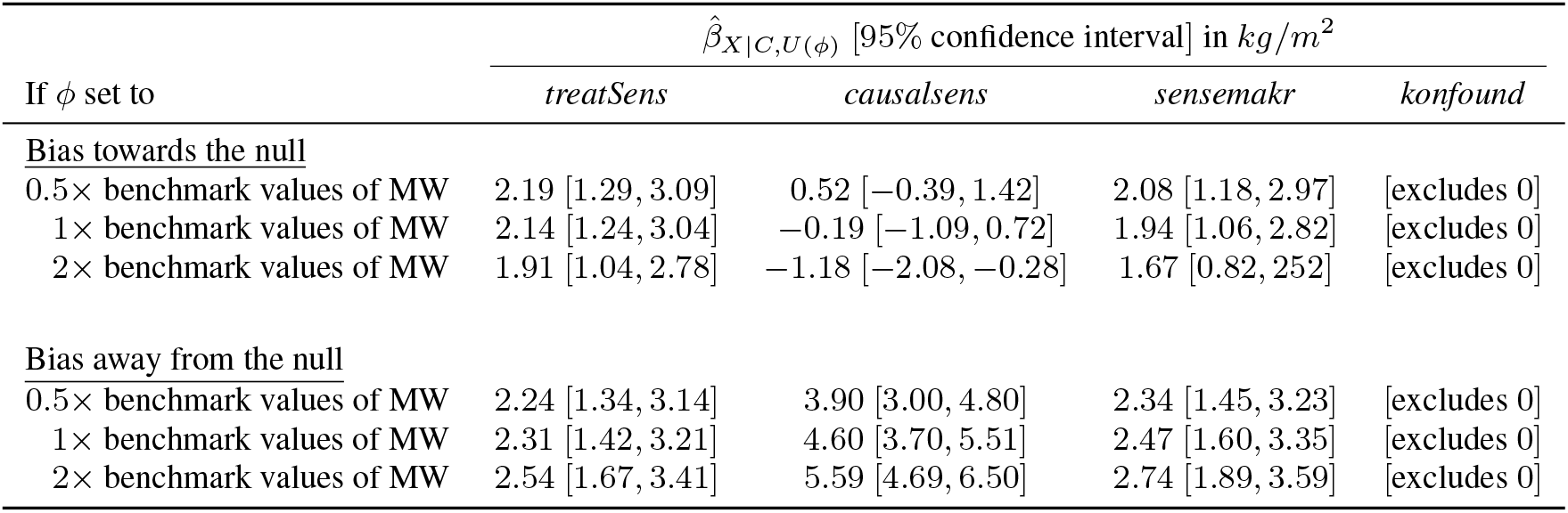
Bias-adjusted estimate 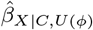 and corresponding 95% confidence interval when bias parameter *ϕ* is set to specified multiples of benchmark values based on strongest measured confounder maternal weight (MW)

Considering unmeasured confounding towards or away from the null, if *U* was comparable to the strongest measured covariate (with respect to its associations with adult BMI and child overweight) then *treatSens* and *sensemakr* report that adjusting for *C* and *U* would give similar results to those of the naive analysis and *konfound* indicates the exposure effect would remain statistically significant. Also, *sensemakr*’s robustness values were substantially higher than the benchmark bounds for 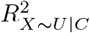 and 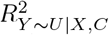 even when these benchmarks were based on all of (Supplementary Table S1). Similarly, the benchmark E-values when omitting the strongest measured covariate and *U* were comparable to the E-values when omitting *U* only (Supplementary Table S2), indicating that the exposure effect adjusted for *C* and *U* would remain above the null and statistically significant. Furthermore, *treatSens, sensemakr*, and *konfound* indicate that *U* would need to be more than double the strength of the strongest measured covariate in order to change the study conclusions (i.e., a null or doubling of the exposure effect, or a statistically insignificant effect). Conversely, *causalsens* suggests adjusting for *U* comparable to the strongest measured covariate could result in an exposure effect close to the null or more than double the naive estimate.

Provided the naive analysis included all of the important confounders then it seems unlikely that the confounding effect of *U*, childhood SEP, could be more than twice as strong as the strongest measured covariate, especially given that childhood SEP would likely be correlated with at least some of the measured covariates. Therefore, under these assumptions, *treatSens, sensemakr, konfound*, and *EValue* indicates robustness of the BCG study conclusions to unmeasured confounding by childhood SEP which was inline with the fully adjusted results. In contrast, *causalsens* suggested study conclusions could differ if we were able to adjust for childhood SEP.

## 6 Discussion

We have conducted an up-to-date review of software implementations of QBA to unmeasured confounding, and a detailed illustration of the latest software applicable for a linear regression analysis of an unmatched study. All programs implement a deterministic QBA, and most are available in the free software environment R. The majority were developed in the latter half of the past decade and include programs available when the naive analysis is a mediation analysis, meta-analysis and a survival analysis. Many programs include features such as benchmarking and graphical displays of the QBA results to aid interpretation. Our comparative example illustrated that even QBA software applicable to the same naive analysis can implement distinct QBA methods. All programs were straightforward to implement and instantly generated the results except for *treatSens* which took about 10 minutes to run when applied to a moderately-sized dataset. All programs provided information about the amount of unmeasured confounding at the tipping points; however, *treatSens, sensemakr* and *causalsens* also provided information on the bias-adjusted results for any specified level of unmeasured confounding with minimal extra burden to the analyst.

Out of the five programs we compared *sensemakr* performs the most detailed QBA. It generates bias-adjusted results for prespecified levels of unmeasured confounding (similarly to *treatSens* and *causalsens*), reports a summary measure at prespecified tipping points (similarly to *EValue* and *konfound*) and conducts a QBA in a worse-case scenario of unmeasured confounding (similarly to *EValue*). However, in our small panel study, three out of seven participants reported difficulties interpreting the output of *sensemakr*. Program *EValue* implements a flexible QBA which can be applied to a wide range of effect measures and makes minimal assumptions about the unmeasured confounding (e.g., allows *U* to be a modifier of the *X* − *Y* relationship). However, the downside of this flexibility is that the analyst may be unaware of the additional assumptions required when converting their effect measure to the risk ratio scale and it can be challenging to establish plausible values for its bias parameters (either from external data or from benchmarking). Also, a notable limitation of programs *EValue* and *konfound* is that they are restricted to establishing robustness to unmeasured confounding (i.e., cannot provide results adjusted for likely levels of unmeasured confounding) and *konfound* only considers sensitivity to changes in statistical significance. The upside of the programs’ simplicity is that they require only summary data and so can be easily applied to multiple published studies, with the *EValue* extended to random-effects meta analyses^51^. Three strengths of *treatSens* over the other programs are: (1) its imputation-style QBA method will be familiar to many analysts, (2) its bias parameters (i.e., regression coefficients) are more likely to be reported by published studies than the bias parameters of the other programs (e.g., partial *R*^2^ values), and (3) *treatSens* can also be applied when the analysis of interest is a non-parametric model (Bayesian additive regression tree). A potential weakness of *treatSens* is that it simulates *U* from a limited choice of joint distributions.

A limitation of our review is that we focused on software described in the published literature. We recognise that additional software programs are available such as other implementations of QBA methods discussed in this review (e.g., another implementation of the E-value^85^) and programs of other QBA methods (e.g., TippingSens^86^). Our illustrative example compared software programs applicable when the analysis of interest is a linear regression since previous comparisons of QBA methods have primarily focused on analyses of binary outcomes^10,19–26^. Of the software we compared, programs *konfound* and *EValue* can be applied to a binary outcome, with *EValue* also applicable when the exposure effect is a hazard ratio. Future work could compare QBA methodology for analyses of other types of outcomes such as survival and categorical outcomes.

Several programs in our review provided benchmark values to aid interpretation of the QBA results. Note that, *sensemakr* can provide benchmark bounds for its bias parameters based on a group of measured covariates which provides a useful aid when considering multiple unmeasured confounders. Interestingly, participants of our small panel study reported difficulties interpreting the E-value in the absence of any benchmarks. One noted issue with benchmarking is that the benchmarks tend to be based on the naive models, *Y* | *X, C* and *X* | *C*, and do not adjust for the omission of *U*^30,63^. See Cinelli and Hazelett for a discussion on why ignoring *U* can affect the benchmark estimates even when *U* is assumed to be independent of *C*^63^. Examples of QBAs using benchmarking that accounts for the omission of *U* include *sensemakr*,^30^, and^87^.

Examples of QBAs tend to focus on a single unmeasured confounder when in fact many weaker unmeasured confounders can jointly change a study’s conclusions^4^. However, several QBA methods are generalisable to multiple unmeasured confounders without burdening the analyst with additional bias parameters. For example, a common assumption is that *U* represents a linear combination of multiple unmeasured confounders, with the elementary scenario that *U* is a single unmeasured confounder. A drawback of this appealing assumption is that the QBA tends to be conservative for multiple unmeasured confounders^63^. Alternatively, a QBA method may leave the functional form of *U* unspecified and instead define its bias parameters as upper bounds (such as the *EValue* where *U* is a categorical variable with categories representing all possible combinations of the multiple unmeasured confounders and its bias parameters *RR*_*XU*_ and *RR*_*UY*_ are the maximum risk ratios comparing any two categories of *U*^78^). A drawback of these upper bounds is that they correspond to extreme situations, making it hard to locate appropriate benchmarks values or external information. To address both drawbacks, a QBA could explicitly model each unmeasured confounder separately whilst allowing for correlations between the confounders, although this would then increase the number of bias parameters. If many unmeasured confounders are suspected, then the analyst should question if a QBA is suitable since the accuracy of a QBA generally relies on a study having measured key confounders. Importantly, a QBA is not a replacement for a correctly designed and conducted study.

In our review, all software implementations were of deterministic QBA methods. In general, deterministic QBA are tipping point analyses with statistical significance as one of the tipping points. Given the call to move away from reliance on statistical significance^88^, we recommend QBA methods that provide bias-adjusted results for all specified values of the bias parameters to give a complete picture of the effect of unmeasured confounding (such as *treatSens, sensemakr* and *causalsens*). However, presenting and interpreting these results can be challenging, especially when there are more than two bias parameters due to the large number of possible value combinations (e.g., three parameters each with 10 possible values gives 1000 combinations). An alternative is a probabilistic QBA which summarises the results as a point estimate and accompanying interval estimate. The advantages of the probabilistic QBA are: (1) the output is familiar to epidemiologists (i.e., similar to point estimate and 95% CI), (2) the interval estimate accounts for all sources of uncertainty due to bias and random sampling, and (3) less reliance on the statistical significance interpretation. Further work is needed to provide software implementations of probabilistic QBAs.

In summary, there have been several new software implementations of QBAs, most of which are available in R. And our comparative evaluation has illustrated the wide diversity in the types of QBA method that can be applied to the same substantive analysis of interest. Such diversity of QBA methods presents challenges in the widespread uptake of QBA methods. Guidelines are needed on the appropriate choice of QBA method, along with provision of software implementations in platforms other than R.

## Data Availability

NHANES study data is publicly available online.
Barry Caerphilly Growth study data is available on request from the study investigator.

## Acknowledgements

We thank the study executives of NHANES, and Dr. P. C. Elwood (MRC Epidemiology Unit, South Wales) and Prof. Y. Ben-Shlomo (University of Bristol) for permitting access to the BCG study data. We also thank researchers at the Leiden University Medical Center for participating in our panel study.

