## Supplementary Materials for "Quantitative bias analysis in practice: Review of software for regression with unmeasured confounding"

#### Supplemental material

Journal Title
XX(X):2–30
©The Author(s) 0000
Reprints and permission:
sagepub.co.uk/journalsPermissions.nav
DOI: 10.1177/ToBeAssigned
www.sagepub.com/
SAGE

### Supplementary Materials

### Quantitative bias analysis in practice: Review of software for regression with unmeasured confounding

**E Kawabata[1,2], K Tilling[1,2], RHH Groenwold[3,4] and RA Hughes[1,2]**

[1]MRC Integrative Epidemiology Unit, University of Bristol, Bristol, United Kingdom
[2]Population Health Sciences, Bristol Medical School, University of Bristol, Bristol, United Kingdom
[3]Department of Clinical Epidemiology, Leiden University Medical Center, Leiden, The Netherlands
[4]Department of Biomedical Data Sciences, Leiden University Medical Center, Leiden, The Netherlands

**Corresponding author:**
Emily Kawabata, Oakfield House, Oakfield Grove, Bristol, BS8 2BN, United Kingdom


**The Barry Caerphilly Growth (BCG) study** **12**

**The National Health and Nutrition Examination Survey (NHANES)** **17**
Description of the study . . . . . . . . . . . . . . . . . . . . . . . . . . . . . . . . 17
Results . . . . . . . . . . . . . . . . . . . . . . . . . . . . . . . . . . . . . . . . 21

**The panel study** **27**

#### Literature search

Box 1: Web of Science search terms for software review. See `https://clarivate.libguides.com/woscc` on how to interpret these terms which are specific to the Web of Science Core Collection.

(TS=(unmeasured NEAR/3 confound*) OR TS=(unmeasured NEAR/3 variable*) OR TS=(unmeasured NEAR/3 covariate*) OR TS=(unmeasured NEAR/3 factor*) OR TS=(unmeasured NEAR/3 predictor*) OR TS=(uncontrolled NEAR/3 confound*) OR TS=(uncontrolled NEAR/3 variable*) OR TS=(uncontrolled NEAR/3 covariate*) OR TS=(uncontrolled NEAR/3 factor*) OR TS=(uncontrolled NEAR/3 predictor*) OR TS=(omitted NEAR/3 confound*) OR TS=(omitted NEAR/3 variable*) OR TS=(omitted NEAR/3 covariate*) OR TS=(omitted NEAR/3 factor*) OR TS=(omitted NEAR/3 predictor*) OR TS=(omission NEAR/3 confound*) OR TS=(unobserved NEAR/3 confound*) OR TS=(unobserved NEAR/3 variable*) OR TS=(unobserved NEAR/3 covariate*) OR TS=(unobserved NEAR/3 factor*) OR TS=(unobserved NEAR/3 predictor*) OR TS=(hidden NEAR/3 confound*) OR TS=(hidden NEAR/3 variable*) OR TS=(hidden NEAR/3 covariate*) OR TS=(hidden NEAR/3 factor*) OR TS=(hidden NEAR/3 predictor*) OR TS=(selection NEAR/3 bias*) OR TS=(residual NEAR/3 confound*) OR TS=(hidden NEAR/3 bias*))
AND
(TS=(sensitivity NEAR/3 analy*) OR TS=(bias NEAR/3 analys*) OR TS=(bias NEAR/3 model*))
AND
(TS=(confound*))

**Figure S1.** Flowchart of the review

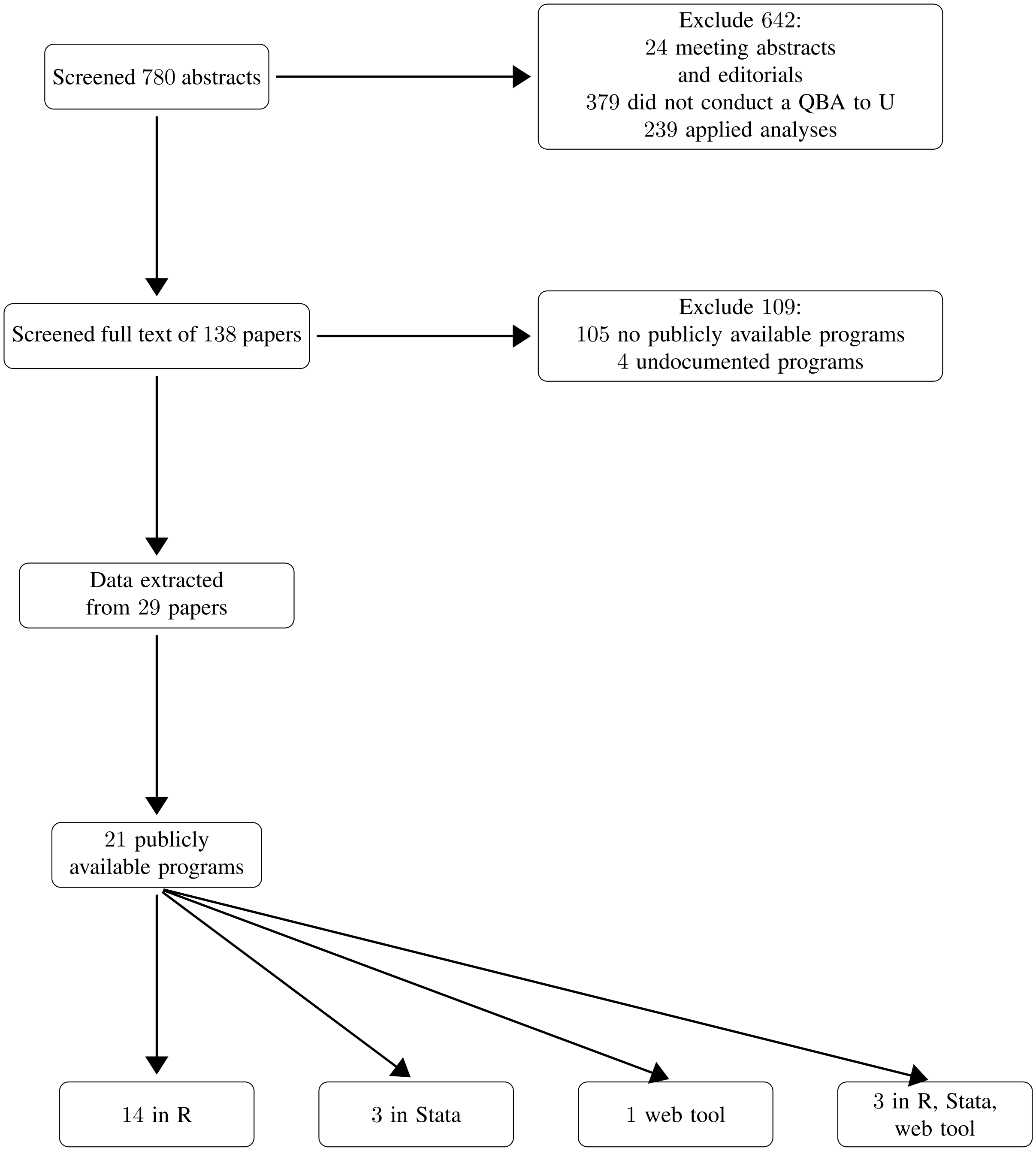

#### Further details on software implementing a quantitative bias analysis for linear regression

In this section we give further details on the five quantitative bias analysis (QBA) programs we discussed in the main text. These programs implement a QBA to unmeasured confounding when the naive analysis is an unmatched analysis, where the exposure is binary and the exposure effect is estimated by a linear regression model.

##### *treatSens*

The QBA method of *treatSens* simulates $U$ under a joint model for the conditional distribution $Y, X, U|C$ and prespecified values of bias parameters $\phi = (\zeta_Z, \zeta_Y)$ and then fits a regression of $Y$ on $X$ adjusting for $C$ and the simulated $U$, recording the exposure coefficient and its standard error[1]. The augmented data are then analysed by a regression of $Y$ on $X$ adjusting for $C$ and the simulated $U$, recording the exposure coefficient and its standard error. This process of simulating $U$ and then analysing the augmented data is repeated $K$ $(\geq 2)$ times and the $K$ sets of exposure estimates and corresponding standard errors $[\hat{\beta}^k_{X|C,U(\phi)}$, $se(\hat{\beta}^k_{X|C,U(\phi)})$ for $k = 1, 2, \ldots, K]$ are combined to generate the bias-adjusted estimate $\hat{\beta}_{X|C,U(\phi)} = \frac{1}{K}\sum_{k=1}^{K}\hat{\beta}^k_{X|C,U(\phi)}$ and its standard error $se(\hat{\beta}_{X|C,U(\phi)}) = \sqrt{W + (1 + K^{-1})B}$ where $W = \frac{1}{K}\sum_{k=1}^{K} se(\hat{\beta}^k_{X|C,U(\phi)})$ reflects the conventional sampling variance of the exposure effect for a given simulated value of $U$, and $B = \frac{1}{K-1}\sum_{k=1}^{K}(\hat{\beta}^k_{X|C,U(\phi)} - (\hat{\beta}_{X|C,U(\phi)})^2)$ reflects the extra variance due to simulating $U$. This whole process is repeated for different prespecified values of $\phi$.

The joint model for $Y, X, U|C$ is the bias model which consists of three sub-models, $p(Y, X, U|C) = p(Y|X, C, U)p(X|C, U)p(U)$: $p(Y|X, C, U)$ is the analysis model which estimates the exposure effect adjusting for measured and unmeasured confounding, $p(X|C, U)$ is the treatment model, and $p(U)$ is a model for the marginal distribution of $U$. The bias model is available as a parametric[1] and semi-parametric model[2]. For the parametric model, the analysis model, $p(Y|X, C, U)$, is a linear regression, and the treatment model, $p(X|C, U)$, is either a linear regression for continuous $X$ or a Probit regression for binary $X$. The semi-parametric model is only available for binary $X$, where $p(Y|X, C, U)$ is a Bayesian Additive Regression Tree (BART) model[3] and $p(X|C, U)$ is either a Probit regression or a BART model. The marginal distribution of $U$, $p(U)$, is normal when $X$ is continuous and Bernoulli when $X$ is binary. Note that, $U$ denotes the part of the unmeasured confounding that is independent of measured covariates $C$ (i.e., $U$ is independent of $C$). Also, $U$ can represent a single unmeasured confounder or a linear combination of multiple (continuous) unmeasured confounders.

$U$ is simulated from the conditional distribution implied by the bias model using an algorithm specific to each of the three types of models (i.e., parametric bias model with continuous $X$, parametric bias model with binary $X$, and semi-parametric bias model with binary $X$) as described in[1,2].

Program *treatSens* is available as an R package from GitHub page `https://github.com/vdorie/treatSens` and it requires individual participant data. The analyst can use *treatSens* when the estimand of interest is the average treatment effect (ATE), the average treatment effect among the treated (ATT), or the average treatment effect among the controls (ATC). Other options include: (1) reparameterising the bias parameters as partial correlations instead of model coefficients (only for continuous $X$), (2) specifying multiple central processing unit cores for parallel processing to help speed-up the run-time of *treatSens*, and (3) specifying the number of times $U$ is simulated for each combination of $\zeta^Y$ and $\zeta^Z$ (default is 20).

#### *causalsens*

Program *causalsens* implements a QBA method proposed by Robins[4,5] which is based on the potential outcomes framework[6]. For binary exposure $X$, let $Y(0)$ and $Y(1)$ denote the potential outcomes to non-exposure and exposure, respectively, and function $f(x, c) = E[Y(x)|X = x, C = c] - E[Y(x)|X = 1 - x, C = c]$ represent the average difference in the potential outcome $Y(x)$ (for $x = 0$ or 1) between the exposure groups, among the subgroup of individuals with $C = c$. Function $f(x, c)$ is called the "confounding function" and setting $f(x, c) = 0$ corresponds to the assumption of no unmeasured confounding, conditional on $C = c$. Examples of the functional form of $f(x, c)$ are given in[7]. Note that, the confounding function is parameterised by a single bias parameter $\alpha$ which implies a combination of the strengths of the $X - U$ and $Y - U$ relationships, conditional on $C$. Since bias parameter $\alpha$ is difficult to interpret, *causalsens* offers the alternative parameterisation, $R^2_\alpha$. See[8] for explanation of how to derive $R^2_\alpha$ from $\alpha$.

Program *causalsens* has two options for $f(x, c)$: the one-sided function $f(x, c) = \alpha(2x - 1)$ and the alignment function $f(X = x, C = c) = \alpha$. The one-sided function assumes the true exposure effect is identical in the exposed and unexposed groups. When $\alpha > 0$ the mean of the potential outcomes to exposure (and non-exposure) is higher for the exposed group than the unexposed group, leading $\hat{\beta}_{X|C}$ to be positively biased; and vice versa for $\alpha < 0$[7]. Under the alignment function the true exposure effect can differ between the exposure groups (i.e., $U$ is an effect modifier). Naive estimate $\hat{\beta}_{X|C}$ is biased by $\alpha(\Pr[X = 0|C = c]Pr[X = 1|C = c])$, where $\Pr[X = 0|C = c]$ and $\Pr[X = 1|C = c]$ denote the probability of being unexposed and exposed, respectively[7]. When the same proportion of people are exposed and unexposed ($\Pr[X = 1|C = c] = \Pr[X = 0|C = c] = 0.5$) then $\hat{\beta}_{X|C}$ is unbiased by $U$. However, when proportionally more individuals are unexposed than exposed (i.e., $\Pr[X = 0|C = c] > \Pr[X = 1|C = c]$) then $\hat{\beta}_{X|C}$ is positively biased for $\alpha > 0$ and negatively biased for $\alpha < 0$; and vice versa when $\Pr[X = 0|C = c] < \Pr[X = 1|C = c]$[7].

Program *causalsens* is an R package available from the Comprehensive R Archive Network (CRAN) or GitHub page `https://github.com/mattblackwell/causalsens` and it requires individual participant data. Exposure $X$ must be binary and outcome $Y$ can be continuous or binary although the naive analysis is restricted to a linear regression. Program *causalsens* can be applied when the estimand of interest is the ATE or ATT (or ATC). The program outputs results with respect to the original bias parameter, $\alpha$, and the alternative parameterisation, $R^2_\alpha$. By default, *causalsens* chooses values for the bias parameter based on the distribution of the outcome, $Y$. The analyst can override this default to specify their own values and can customise their own confounding function.

The output of *causalsens* does not explicitly indicate the tipping points of the point estimate or CI. Instead using the outputted plot, the analyst can determine the values of bias parameter, $\phi$, at which the bias-adjusted estimate, $\hat{\beta}_{X|C,U(\phi)}$, equals a specific value (e.g., the null) and the CI includes the null. Note that, the R package downloaded from CRAN fixes the statistical significance to be at the $5\%$ level whereas the R package downloaded from GitHub allows the analyst to change the statistical significance level.

#### *sensemakr*

The absolute difference between the naive and bias-adjusted estimates, given values of $\phi = (R^2_{X\sim U|C}, R^2_{Y\sim U|X,C})$ is calculated as

$$|\widehat{\Delta}_\phi| = se(\hat{\beta}_{X|C})\sqrt{\frac{R^2_{Y\sim U|X,C}R^2_{X\sim U|C}}{1-R^2_{X\sim U|C}} \times df}, \tag{1}$$

where $se(\hat{\beta}_{X|C})$ and *df* are the standard error of the exposure effect and degrees of freedom from the naive analysis, respectively. Depending on the direction of effect of $U$, $\widehat{\Delta}_\phi$ is either added to or subtracted from $\hat{\beta}_{X|C}$ to obtain $\hat{\beta}_{X|C,U(\phi)}$ and its standard error is calculated as

$$se(\hat{\beta}_{X|C,U(\phi)}) = se(\hat{\beta}_{X|C})\sqrt{\frac{1-R^2_{Y\sim U|X,C}}{1-R^2_{X\sim U|C}} \times \frac{df}{df-1}}. \tag{2}$$

The robustness value of the point estimate can be calculated by setting $R^2_{X\sim U|C} = R^2_{Y\sim U|X,C}$ and $\widehat{\Delta}_\phi$ to the relevant tipping point value, such as $\hat{\beta}_{X|C}$ for a null effect, and then solving equation (1) for $R^2_{X\sim U|C}$. Similarly for the t-value, where we first calculate $se(\hat{\beta}_{X|C,U(\phi)})$ using equation (2) (which simplifies to a function of $se(\hat{\beta}_{X|C})$ and *df*) and then derive the value of $\widehat{\Delta}_\phi$ which corresponds to a $\kappa\%$ critical value of the Student's t-distribution with *df* degrees of freedom (for a given null or non-null hypothesis).

Program *sensemakr* makes no distributional assumptions about $U$ but does assume that $U$ is either a single unmeasured confounder or a linear combination of two or more unmeasured confounders. Cinelli and Hazlett state that their assumption about multiple unmeasured confounders is conservative[9].

Program *sensemakr* also implements a QBA for an extreme scenario which is a worst-case setting where all of the unexplained variation in outcome $Y$ is due to $U$ (i.e., $R^2_{Y\sim U|X,C} = 1$). For this extreme scenario, the program outputs $(1)$ summary measures for the point estimate and t-value of the minimum strength of the relationship between $X$ and $U$ (i.e., $R^2_{X\sim U|C}$) in order to change the study conclusions, $(2)$ benchmark bounds for $R^2_{X\sim U|C}$, and $(3)$ a plot of the bias-adjusted results for different values of $R^2_{X\sim U|C}$. Note that, the R package will also generate the plot for different extreme scenarios such as $R^2_{Y\sim U|X,C} = 0.75$.

Program *sensemakr* is available as an R package (install from CRAN or github page `https://github.com/carloscinelli/sensemakr`), Stata command (install from the Statistical Software Components (SSC) archive) and as a Shiny app (`https://carloscinelli.shinyapps.io/robustness_value/`). The analyst can either apply *sensemakr* to their individual participant data using R or Stata or input summary data from the naive analysis using R or the web tool. By default, the direction of the effect of $U$ is towards the null and the tipping points for the point estimate and t-value are the null and t-critical value at $5\%$ statistical significance, respectively. Available options allow the analyst to set the direction of effect to be away from the null, a different statistical significance level, and a non-null value for the tipping point. All three implementations will generate results under the extreme scenario.

#### *EValue*

The rationale of the E-value is based on an upper bound, $BF_\phi$, for the maximum amount of bias due to unmeasured confounding that will reduce a naive risk ratio (e.g., point estimate of the exposure effect adjusted for $C$ only) to a prespecified level (e.g., the null)[10,11]. The upper bound, $BF_\phi$, is defined on the risk ratio scale as

$$BF_\phi = \frac{RR_{UY} RR_{XU}}{RR_{UY} + RR_{XU} - 1}, \tag{3}$$

where $RR_{UY}$ and $RR_{XU}$ are the bias parameters for the $Y - U$ and $X - U$ associations, respectively. Assuming the naive risk ratio, $\widehat{RR}$, is greater than 1 then the E-value for reducing $\widehat{RR}$ to at least the null (i.e., $\leq 1$) is the minimum value of the bias parameters when $RR_{UY}$ and $RR_{XU}$ are equal to each other and $BF_\phi$ equals $\widehat{RR}$. The formula for the E-value is

$$\text{E-value } = \widehat{RR} + \sqrt{\widehat{RR} \times (\widehat{RR} - 1)} \tag{4}$$

and can be derived from equation (3) by setting $BF_\phi = \widehat{RR}$ and E-value $= RR_{UY} = RR_{XU}$ and then solving the resulting quadratic equation. When the naive risk ratio is less than 1, equation (4) is applied with $\widehat{RR}$ set to the inverse of the naive risk ratio. See[12] for further technical details of the E-value. When the naive exposure effect, $\hat{\beta}_{X|C}$, is a risk ratio then the E-value for a null exposure effect is calculated by setting $\widehat{RR}$ equal to $\hat{\beta}_{X|C}$. Similarly, the E-value for a statistically insignificant exposure effect is calculated by setting $\widehat{RR}$ equal to the CI limit of $\hat{\beta}_{X|C}$ closest to the null. When the exposure effect is not a risk ratio (e.g., a mean difference, risk difference, odds ratio or hazard ratio) then $\hat{\beta}_{X|C}$ and its CI limit are first converted to the risk ratio scale before applying equation (4)[11]. Note that, the E-value can also be calculated when the tipping point for the point estimate is a non-null value such as the maximum value of the exposure effect that is clinically irrelevant (see Supplementary Materials of[11] for more details).

The E-value makes no distributional assumptions about the unmeasured confounding, where $U$ can denote a single or multiple unmeasured confounders of disparate variable types[10]. Also, the E-value allows $U$ to be a modifier of the exposure effect. However, for effect measures other than the risk ratio then additional assumptions are required to convert the naive results to the risk ratio scale[11].

The E-value evaluates the sensitivity to unmeasured confounding in the worst-case scenario[13]. For example, when $U$ is a single, binary unmeasured confounder then the E-value assumes that the prevalence of $U$ is $100\%$ in one of the two exposure groups[14]. Therefore, the E-value is limited to establishing robustness to unmeasured confounding.

Software to calculate an E-value is available in R (package *EValue*[15,16] available from CRAN), in Stata (command *evalue*[17] available from the SSC archive and from GitHub page `https://github.com/mayamathur/evalue_package`) and as a web tool (the *E-value calculator*[15] available at `https://www.evalue-calculator.com/evalue`). All implementations only require summary data (e.g., point estimate and CI limit from the naive analysis) and output E-values for the point estimate and CI limit. Additionally, the online tool, the *E-value Calculator*, includes a feature that outputs the bias factor, $BF_\phi$, and corresponding bias-adjusted estimate of the point estimate or CI limit (on the risk ratio scale) for prespecified values of $RR_{UY}$ and $RR_{XU}$.

The analyst can specify a null or non-null value for the tipping point of the point estimate (null is the default setting). For effect measures other than the risk difference and mean difference, the analyst inputs the naive point estimate and CI limit closest to the null and so the level of statistical significance is automatically set by the analyst. For a risk difference, the analyst inputs the cells of a $2 \times 2$ table (i.e., number of exposed individuals who experienced the outcome, number of unexposed individuals who experienced the outcome, number of exposed individuals who did not experience the outcome, and the number of unexposed individuals who did not experience the outcome) and can specify the statistical significance level (5% is the default setting). For a mean difference, the analyst inputs a standardised mean difference and corresponding standard error but the statistical significance is fixed at 5%.

In addition to computing E-values for unmeasured confounding in a single study, the *E-value Calculator* and R package *EValue* include features to calculate an E-value to assess sensitivity to unmeasured confounding in a meta-analysis[18] and an E-value to assess sensitivity to selection bias[19]. Also, R package *EValue* can implement a sensitivity analysis to differential misclassification[20], multiple sources of bias simultaneously (i.e., confounding, selection bias, and misclassification)[21], and assess sensitivity of effect heterogeneity estimates to unmeasured confounding[22].

#### *konfound*

*konfound* reports two measures that quantify the level of unmeasured confounding necessary to invalidate or sustain statistical significance of the exposure effect: percent bias and impact threshold.

For statistically significant naive estimate $\hat{\beta}_{X|C}$, standard error $se(\hat{\beta}_{X|C})$ and degrees of freedom *df*, the percent bias to invalidate inference is calculated as

$$\text{percent bias } = 100 \times \left(1 - \frac{\beta^{\kappa}_{X|C,U}}{\hat{\beta}_{X|C}}\right),$$

where $\beta^{\kappa}_{X|C,U}$ is the threshold value of the exposure effect for $\kappa\%$ statistical significance (i.e., the two-sided P-value of $\beta^{\kappa}_{X|C,U}$ is exactly $\kappa\%$)[23]. Note that, $\beta^{\kappa}_{X|C,U} = sgn(\hat{\beta}_{X|C}) \times se(\hat{\beta}_{X|C}) \times t_{\kappa,df}$ where $sgn(\hat{\beta}_{X|C})$ and $se(\hat{\beta}_{X|C})$ are the sign and standard error of $\hat{\beta}_{X|C}$, respectively, and $t_{\kappa,df}$ is the $\kappa\%$ critical value of the Student's $t$-distribution with *df* degrees of freedom. For example, for $\hat{\beta}_{X|C} = 2$, $se(\hat{\beta}_{X|C}) = 0.4$ and 95 degrees of freedom, the value of $\beta^{\kappa}_{X|C,U}$ corresponding to 5% statistical significance is $0.4 \times 1.985 = 0.794$ which gives a percent bias of 60.3%. Therefore, inference would be invalidated if the magnitude of the bias-adjusted estimate of the exposure effect was less than 0.794 (i.e., $|\hat{\beta}_{X|C,U(\phi)}| < 0.794$).

The impact threshold is based on a partial correlation framework[24]. The partial correlation between $X$ and $Y$ given $C$ and $U$, $r_{Y\sim X|C,U(\phi)}$, can be written as follows

$$r_{Y\sim X|C,U(\phi)} = \frac{r_{Y\sim X|C} - r_{Y\sim U|C} \times r_{X\sim U|C}}{\sqrt{1 - r^2_{Y\sim U|C}}\sqrt{1 - r^2_{X\sim U|C}}}, \tag{5}$$

where $r_{Y\sim X|C}$ is the partial correlation between $X$ and $Y$ given $C$ from the naive analysis, and $r_{X\sim U|C}$ and $r_{Y\sim U|C}$ are the two bias parameters[23]. The impact threshold is the product of the minimum value of $r_{X\sim U|C}$ and $r_{Y\sim U|C}$, when $|r_{Y\sim U|C}| = |r_{X\sim U|C}|$, such that $r_{Y\sim X|C,U(\phi)}$ equals $r^{\kappa}_{Y\sim X|C,U}$, the threshold value for the partial correlation between $X$ and $Y$ given $C$ and $U$ when the two-sided P-value for the exposure effect is exactly $\kappa\%$. Value $r^{\kappa}_{Y\sim X|C,U}$ is given by

$$r^{\kappa}_{Y\sim X|C,U} = sgn(r_{Y\sim X|C}) \times \frac{t_{\kappa,df}}{\sqrt{n-q-1+t^2_{\kappa,df}}},$$

where $sgn(r_{Y\sim X|C})$ denotes the sign (or direction) of $r_{Y\sim X|C}$, $n$ is the sample size, $q$ is the number of independent variables (i.e., $X$ and $C$), and $t_{\kappa,df}$ is the $\kappa\%$ critical value of the Student's $t$-distribution with *df* degrees of freedom. Setting $r_{Y\sim X|C,U(\phi)} = r^{\kappa}_{Y\sim X|C,U}$, $|r_{Y\sim U|C}| = |r_{X\sim U|C}|$, and impact $= r_{Y\sim U|C} \times r_{X\sim U|C}$ then equation (5) can be rearranged as follows

$$\text{impact} = \frac{r_{Y\sim X|C} - r^{\kappa}_{Y\sim X|C,U}}{1-|r^{\kappa}_{Y\sim X|C,U}|}.$$

When $r_{Y\sim X|C} > r_{Y\sim X|C,U}$ the impact threshold is calculated as[23]

$$\text{impact} = \frac{r_{Y\sim X|C} - r^{\kappa}_{Y\sim X|C,U}}{1-r^{\kappa}_{Y\sim X|C,U}}.$$

And, when $r_{Y\sim X|C} < r_{Y\sim X|C,U}$ the impact threshold is calculated as

$$\text{impact} = \frac{r_{Y\sim X|C} - r^{\kappa}_{Y\sim X|C,U}}{1+r^{\kappa}_{Y\sim X|C,U}}.$$

For example, let $\hat{\beta}_{X|C} = 2$, $se(\hat{\beta}_{X|C}) = 0.4$, $df = 95$ and $r_{Y\sim X|C} = 0.456$. At $5\%$ statistical significance, $t_{\kappa,df} = 1.985$, the threshold value, $r^{\kappa}_{Y\sim X|C,U}$, is $0.200$, and impact $= 0.321$. Therefore, the magnitudes of the partial correlations of $U$ with $X$ and with $Y$ given $C$ must both exceed $0.567$ in order to invalidate an inference for a null hypothesis at the $5\%$ level.

Software *konfound* is available as an R package (from CRAN or GitHub page `https://github.com/jrosen48/konfound`), a Stata command (see[23] for download instructions), and a web tool (Shiny app `https://jmichaelrosenberg.shinyapps.io/konfound-it/`). The R and Stata implementations can be applied to individual participant data and to summary data (i.e., point estimate, standard error, sample size and number of measured covariates from the naive analysis) using function or command *pkonfound*. The web tool and R package can be used with two types of summary data: (1) point estimate, standard error, sample size and number of measured covariates from the naive analysis, and (2) $2 \times 2$ cross tabulation of exposure and outcome data.

Note that, the Stata command only generates a threshold plot to assess sensitivity to an invalidated inference (i.e., when $\hat{\beta}_{X|C}$ is statistically significant) whereas the R package and online tool will generate a threshold plot to assess sensitivity to an invalidated or sustained inference (i.e., when $\hat{\beta}_{X|C}$ is statistically significant and insignificant). By default, all implementations assume the significance level is $5\%$ and the null hypothesis is "no exposure effect". These settings can be changed in the R and Stata implementations. Other options include application of the QBA to multiple studies (function or command *mkonfound* in R and Stata, respectively) and a nonlinear option for application when the analysis model is a logistic or probit regression, although the R package and Stata command reports a warning to ignore the impact threshold for a binary outcome.

#### The Barry Caerphilly Growth (BCG) study

In this section, additional results of the QBA analysis of the Barry Caerphilly Growth study are presented.

**Figure S2.** Quantitative bias analysis for effect of child overweight on adult body mass index from the Barry Caerphilly Growth study. *sensemakr* contour plots when unmeasured confounding increases the exposure effect: black contours (bias-adjusted estimates for the point estimate in (a) and t-value at $5\%$ significance in (b)), diamonds (benchmarks), and black triangle (naive estimate).

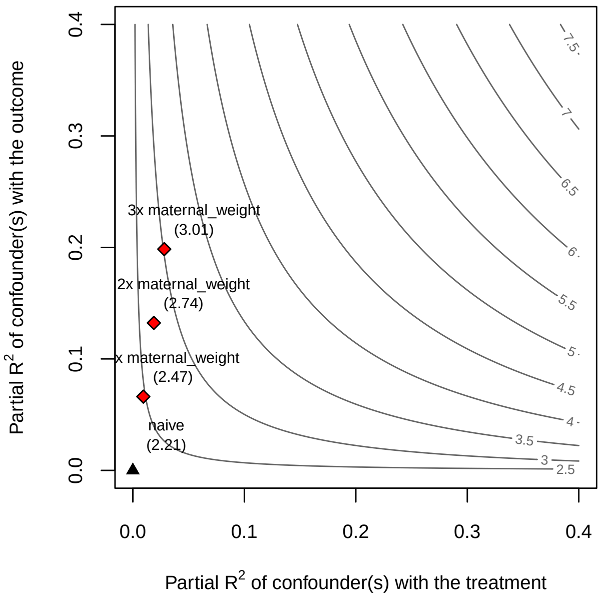

**(a)** *sensemakr* for point estimate

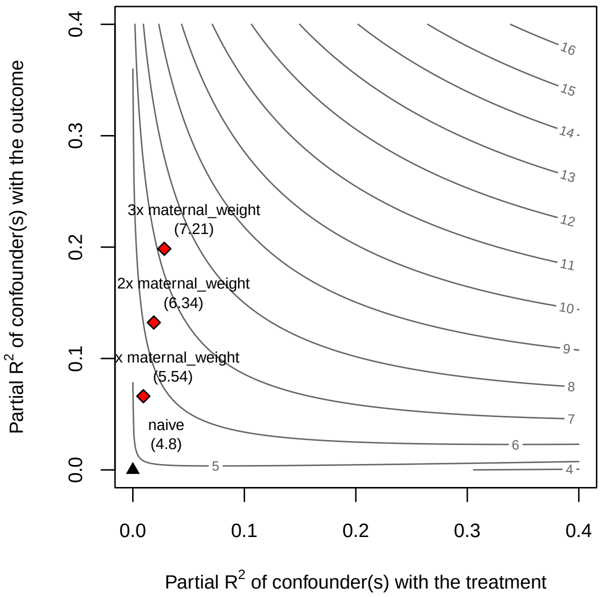

**(b)** *sensemakr* for t-value

**Table S1.** Benchmark upper bounds for sensemakr's bias parameters, $R^2_{X\sim U|C}$ and $R^2_{Y\sim U|X,C}$, based on partial $R^2$ of a measured covariate (or group of covariates) with exposure $X$ and outcome $Y$. Data from the Barry Caerphilly Growth study.

| Measured covariate | $R^2_{X\sim U\|C}(\%)$ | $R^2_{Y\sim U\|X,C}(\%)$ |
|---|---|---|
| Sex | 0.89 | 0.00 |
| Gestational age | 0.10 | 0.14 |
| Birth weight | 2.52 | 0.07 |
| Paternal height | 0.14 | 1.03 |
| Paternal weight | 0.73 | 3.77 |
| Maternal height | 0.27 | 1.43 |
| Maternal weight | 0.94 | 6.61 |
| All covariates | 5.47 | 13.52 |

**Figure S3.** E-value quantitative bias analysis plot for the effect of childhood overweight on adult body mass index from the Barry Caerphilly Growth study. Curves denote value combinations of the bias parameters that explain away $100\%$ of the naive point estimate (red) and statistical significance at the $5\%$ level (black); dots denote corresponding E-value.

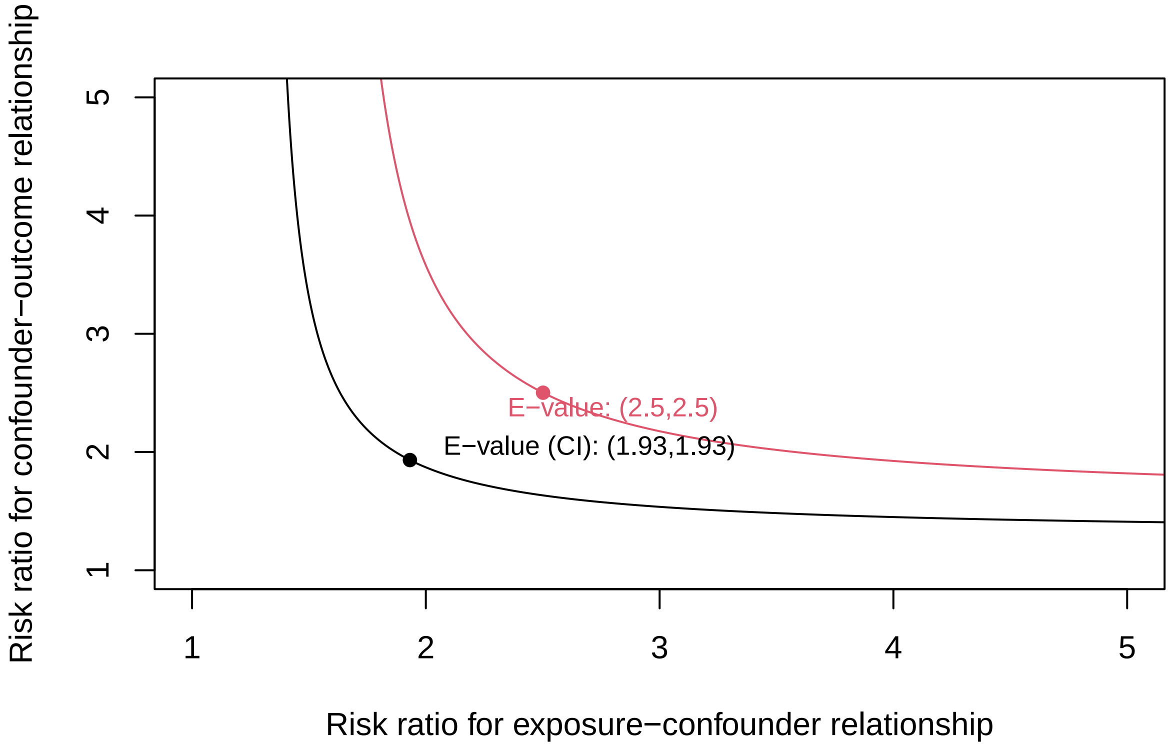

**Table S2.** Benchmark E-values, for the point estimate and lower confidence interval (CI) limit, based on omitting each measured covariate while controlling for the remaining covariates. For the effect of child overweight on adult body mass index from the Barry Caerphilly Growth study.

| | E-value | |
|---|---|---|
| Measured covariate | Point estimate | Lower CI limit |
| Sex | 2.50 | 1.94 |
| Gestational age | 2.49 | 1.92 |
| Birth weight | 2.53 | 1.96 |
| Paternal height | 2.53 | 1.95 |
| Paternal weight | 2.62 | 2.03 |
| Maternal height | 2.55 | 1.97 |
| Maternal weight | 2.68 | 2.08 |

**Figure S4.** *konfound*: Threshold plot representing the percent bias necessary to invalidate inference (at $5\%$ statistical significance) for the effect of childhood obesity on adult body mass index from the Barry Caerphilly Growth study.

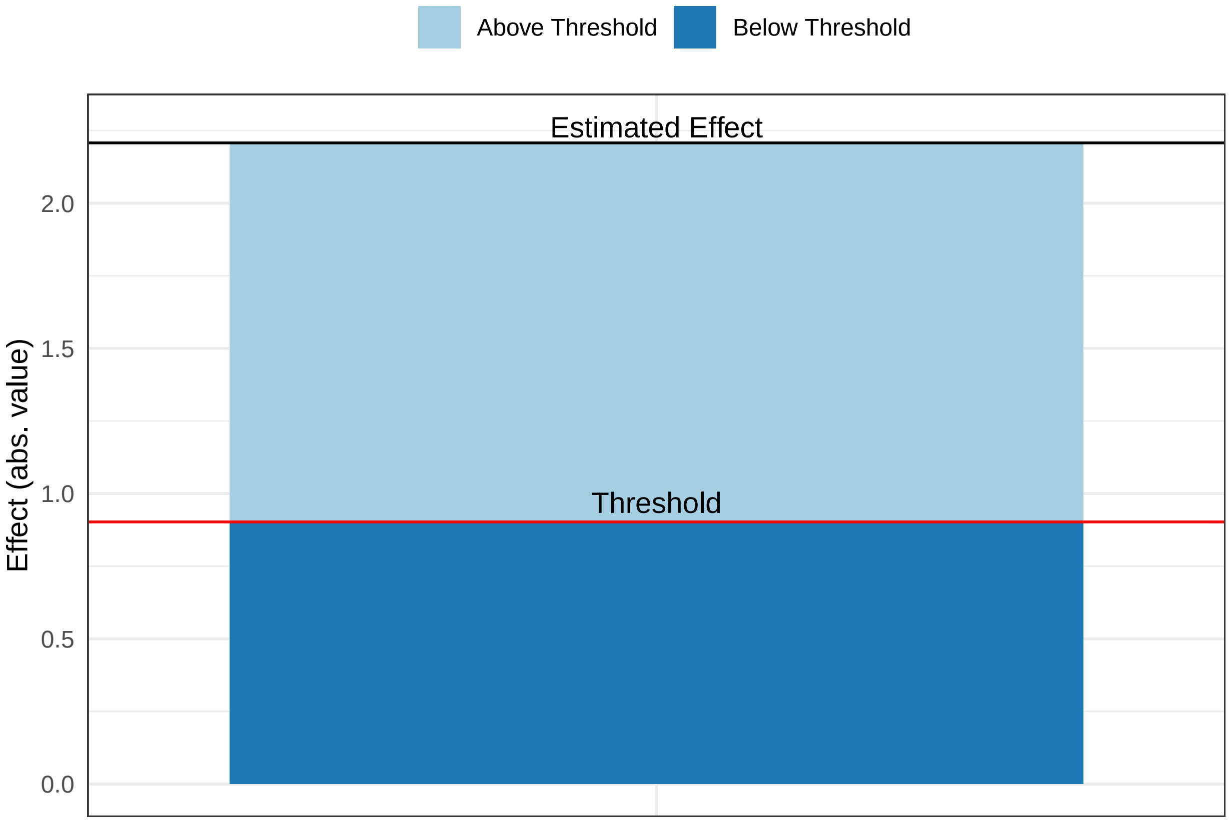

**Figure S5.** *konfound*: Correlation plot of the impact of unmeasured confounding necessary to invalidate inference (at $5\%$ statistical significance) for the effect of childhood body obesity on adult body mass index from the Barry Caerphilly Growth study. Partial correlations $R_{x \cdot cv|Z}$ and $R_{y \cdot cv|Z}$ are between unmeasured confounder $cv$ and exposure $X$, and outcome $Y$, respectively, after accounting for measured covariates $Z$.

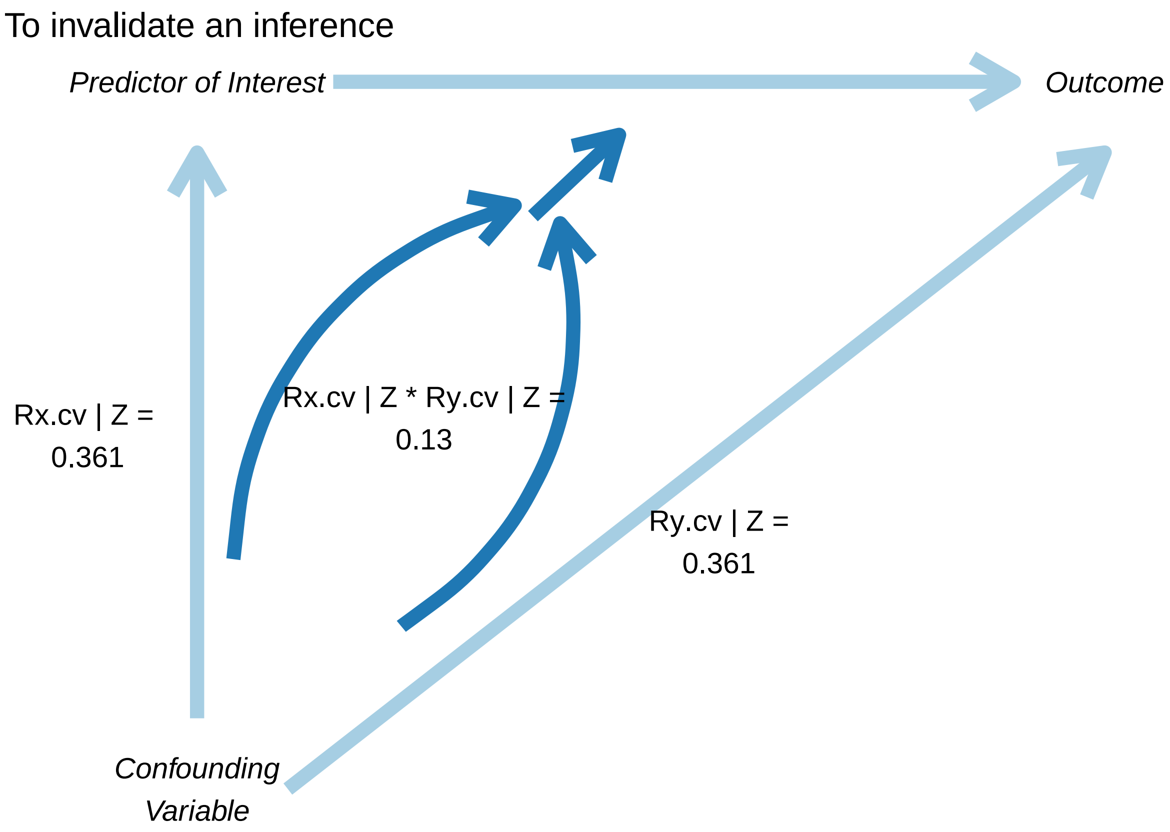

**Table S3.** *konfound*: Benchmark values for $r_{X\sim U|C}$ and $r_{Y\sim U|C}$, and impact threshold $r_{X\sim U|C} \times r_{Y\sim U|C}$, based on partial correlations of each measured covariate with childhood obesity and adult body mass index (BMI). Data from the Barry Caerphilly Growth study.

| Measured covariate | Partial correlation with childhood obesity | Partial correlation with adult BMI | Impact threshold |
|---|---|---|---|
| Sex | $0.0938$ | $0.0216$ | $0.0020$ |
| Gestational age | $-0.0312$ | $0.0298$ | $-0.0009$ |
| Birth weight | $0.1568$ | $0.0576$ | $0.0090$ |
| Paternal height | $-0.0368$ | $-0.1062$ | $0.0039$ |
| Paternal weight | $0.0854$ | $0.2030$ | $0.0173$ |
| Maternal height | $-0.0518$ | $-0.1267$ | $0.0066$ |
| Maternal weight | $0.0965$ | $0.2614$ | $0.0252$ |

#### The National Health and Nutrition Examination Survey (NHANES)

We applied *treatSens,causalsens,sensemakr,EValue* and *konfound* to data from the NHANES study. As per the analysis of the BCG study, we used measured variables to represent the unmeasured confounders $U$. So, in effect our analyses examined the effect of not including certain confounders and we assumed that after adjustment for $U$ and $C$ there was no unmeasured confounding.

For *treatSens* we used Probit regression for its treatment model because $X$ was binary, and for *causalsens* we used the one-sided confounding function because we assumed the exposure effect was the same in both exposure groups.

As this is an illustrative example of applying a QBA to unmeasured confounding, we have ignored other potential sources of bias (such as missing data) and only considered a small number of measured covariates. We restricted our analyses to participants with complete data on $Y, X, C$ and $U$.

##### *Description of the study*

The NHANES study consists of a series of health and nutrition surveys conducted by the National Center for Health Statistics. Every year since 1999, approximately $5,000$ individuals of all ages are interviewed in their homes with health examinations conducted in a mobile examination centres. We analysed data from the $2015-2016$ NHANES survey[25].

Our analysis was a linear regression of systolic blood pressure (SBP) on diabetes among adults aged $\geq 18$ years. Diabetes was defined as a HbA1c measurement of at least $6.5\%$ (diabetes$=1$ if HbA1C $\geq 6.5\%$, $0$ otherwise)[26]. Measured covariates $C$ were age and sex, with age as the strongest measured covariate (i.e., largest associations with diabetes and SBP). The unmeasured confounders $U$ were BMI, ethnicity and poverty income ratio (PIR; the ratio of family income to the federal poverty line[27]). Based on the $4,576$ participants with complete data on all variables, $\hat{\beta}_{X|C}$ was $3.48$ mmHg ($99\%$ CI $1.55, 5.40$ mmHg; P-value $< 0.0001$) and the fully adjusted estimate (i.e., adjusted for $C$ and $U$) was $1.67$ mmHg ($99\%$ CI $-0.27, 3.61$ mmHg; P-value $0.03$). So, controlling for BMI, ethnicity and PIR explained $48\%$ of $\hat{\beta}_{X|C}$ and resulted in a $99\%$

CI that contained the null (i.e., P-value greater than $0.01$). Statistical significance was defined at the $1\%$ level.

Note that, *EValue* and the CRAN version of *causalsens* fix the statistical significance at the $5\%$ level. To get around this we used the Github version of *causalsens* (which allows the analyst to change the statistical significance level) and for *EValue* we converted the mean difference (of the point estimate and CI limit closest to the null) to an approximate risk ratio and then used the function applicable when the effect measure is a risk ratio.

We begin with a description of the outputted results and then compare the results across the five programs.

**Figure S6.** Quantitative bias analysis for effect of diabetes on systolic blood pressure from the National Health and Nutrition Examination Survey. Red contour (null effect in (a) and (c), t-value at $1\%$ significance in (d)), blue contours (bracket $1\%$ statistically insignificant estimates), black contour or line (bias-adjusted estimates), grey shaded area ($99\%$ confidence intervals for bias-adjusted estimates), pluses, inverted triangles, crosses, and diamonds (benchmark), and black triangle (naive estimate).

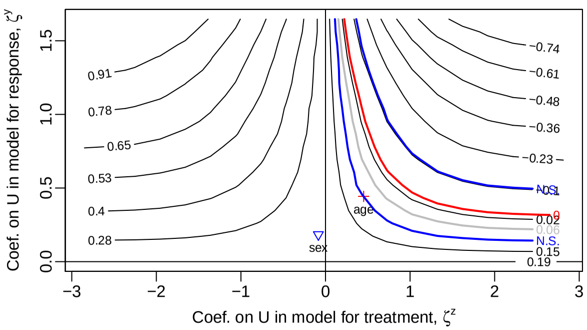

**(a)** *treatSens*

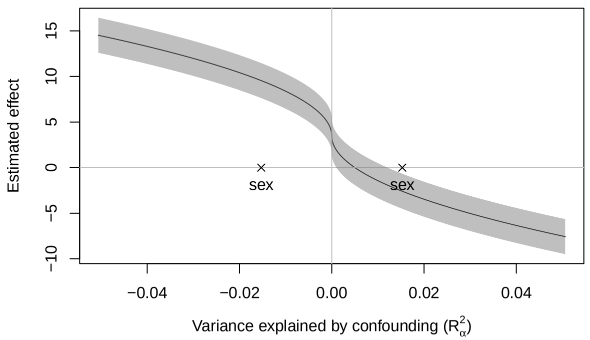

**(b)** *causalsens* (×: sex)

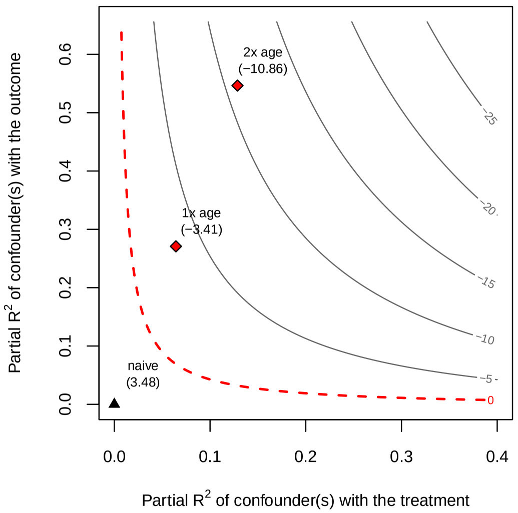

**(c)** *sensemakr* for point estimate

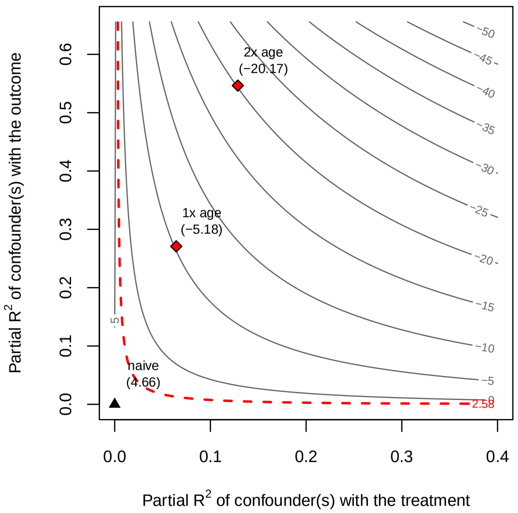

**(d)** *sensemakr* for t-value

#### Results

*treatSens*

Supplementary Figure S6(a) shows the results of the *treatSens* QBA. If the magnitudes of the diabetes$-U$ and SBP-$U$ associations were comparable to those of the strongest measured covariate, age (diabetes$-$age$= 0.45$, and SBP$-$age$= 0.44$ on the standardised scale) then $\hat{\beta}_{X|C,U(\phi)}$ could be $\approx 0.28$ standard deviations of SBP (i.e., $46\%$ increase of $\hat{\beta}_{X|C}$ and statistically significant) or approximately $0.10$ standard deviations of SBP ($48\%$ reduction of $\hat{\beta}_{X|C}$) with a P-value of $0.01$. Potentially, a $U$ comparable to age could explain away the statistical significance of $\hat{\beta}_{X|C}$. For unmeasured confounding to explain away all of $\hat{\beta}_{X|C}$ then $U$ would need to have stronger associations with either diabetes, SBP or both (e.g., double that of diabetes$-$age ($\zeta^z \approx 1, \zeta^y \approx 0.49$), or double that of SBP$-$age ($\zeta^z \approx 0.45, \zeta^y \approx 1$), or in-between for both ($\zeta^z \approx 0.60, \zeta^y \approx 0.75$)).

*causalsens*

Supplementary Figure S6(b) shows the results of the *causalsens* QBA, where $R^2_\alpha > 0$ corresponds to individuals in the diabetic group tending to have higher potential SBP values (to both exposure to diabetes and no exposure) than the non-diabetic group (i.e., individuals in the non-diabetic group tended to be healthier regardless of diabetic status); and the converse for $R^2_\alpha < 0$. Note that, the default scale for $R^2_\alpha > 0$ excluded the benchmark for the strongest measured covariate (age).

If the residual variance explained by $U$ was comparable to that of the weakest measured covariate, sex, ($|R^2_\alpha| = 0.015$ or $1.5\%$) then $\hat{\beta}_{X|C,U(\phi)}$ could be as large as 10 mmHg or a reversed effect of about $-2.5$ mmHg; both with a P-value $< 0.01$. Additionally, if $U$ had a partial $R^2$ value closer to that of age then $\hat{\beta}_{X|C,U(\phi)}$ could be $\leq -10$ mmHg or $\geq 15$ mmHg. A $U$ weaker than covariate sex (with respect to proportion of the explained residual variance) could explain away the statistical significance of $\hat{\beta}_{X|C}$. Furthermore, if individuals in the non-diabetic group were healthier than the diabetic group (regardless of diabetes status; i.e., $R^2_\alpha \approx 0.01$) then $U$ could explain away all of $\hat{\beta}_{X|C}$ (i.e., $\hat{\beta}_{X|C,U(\phi)} = 0$).

*sensemakr*

The robustness values for $\hat{\beta}_{X|C}$ and $1\%$ statistical significance were $6.65\%$ and $3.03\%$, respectively. Values for $R^2_{X\sim U|C}$ and $R^2_{Y\sim U|X,C}$ exceeding $6.65\%$ are plausible given the benchmark bounds reported in Supplementary Table S4. Therefore, we cannot exclude the possibility that unmeasured confounding could explain away all of $\hat{\beta}_{X|C}$ or all of its $1\%$ statistical significance. This is supported by Supplementary Figures S6(c) and (d) which show that even if $U$ was a weaker confounder than age, provided the direction of its effect was to reduce the point estimate, then accounting for $U$ could result in a null or statistically insignificant exposure effect. Depending on the direction of the effect of $U$, if the magnitude of the confounding effect of $U$ was comparable to that of age then the exposure effect could be reversed with $\hat{\beta}_{X|C,U(\phi)} = -3.41$ mmHg (Figure S6(c)) or increased to $\hat{\beta}_{X|C,U(\phi)} = 10.36$ mmHg (Supplementary Figure S7(a)).

*EValue*

The E-values for $\hat{\beta}_{X|C}$ and its $99\%$ lower CI limit were $1.67$ and $1.38$, respectively (Supplementary Figure S8). Supplementary Table S5 reports the benchmark E-values after excluding age and sex, separately. Adjusting for age reduces the E-value for the point estimate and lower CI limit by about $1$ (e.g., moved from $2.61$ to $1.67$). Therefore, if the confounding effect of $U$ was comparable to age, adjusting for age, sex and $U$ could result in E-values close to 1. Therefore, we cannot exclude the possibility that unmeasured confounding could explain away all of $\hat{\beta}_{X|C}$ or all of its $1\%$ statistical significance.

*konfound*

The percent bias and impact threshold were $44.67\%$ and $0.032$, respectively. Therefore, adjusting for $U$ could result in a statistically insignificant exposure effect if $U$ explained away at least $44.67\%$ of $\hat{\beta}_{X|C}$ (i.e., $\hat{\beta}_{X|C,U(\phi)} < 1.93$ mmHg; Supplementary Figure S9) or if the partial correlations of $U$ with SBP and diabetes both exceeded $0.178$ (Supplementary Figure S10). Note that, the benchmark values for $r_{X\sim U|C}$ and $r_{Y\sim U|C}$ based on age (Supplementary Table S6) were larger than $0.178$ implying that a $U$ comparable to age could explain away the statistical significance of the exposure effect.

Comparison of the results

If $U$ was comparable to the strongest measured covariate, age, then *causalsens*, *sensemakr*, and *EValue* indicated that the exposure effect adjusted for $C$ and $U$ would either be null or in the reverse direction, while *treatSens* suggested that the exposure effect would still be positive although not statistically significant at the $1\%$ level. Program *konfound* also indicated that the statistical significance of the exposure effect was not robust to unmeasured confounding (if $U$ was comparable to age). Given there were only two measured covariates, it seems plausible that there could be a $U$ comparable to age (with respect to confounding of the diabetes-SBP relationship) and possibly that there were multiple unmeasured confounders. In context of multiple unmeasured confounders, then the results of *treatSens* also indicate that the exposure effect adjusted for $C$ and $U$ could either be null or in the reverse direction.

**Figure S7.** Quantitative bias analysis for effect of diabetes on systolic blood pressure from the National Health and Nutrition Examination Survey. *sensemakr* contour plots when unmeasured confounding increases the exposure effect: black contours (bias-adjusted estimates for the point estimate in (a) and t-value at $1\%$ significance in (b)), diamonds (benchmarks), and black triangle (naive estimate).

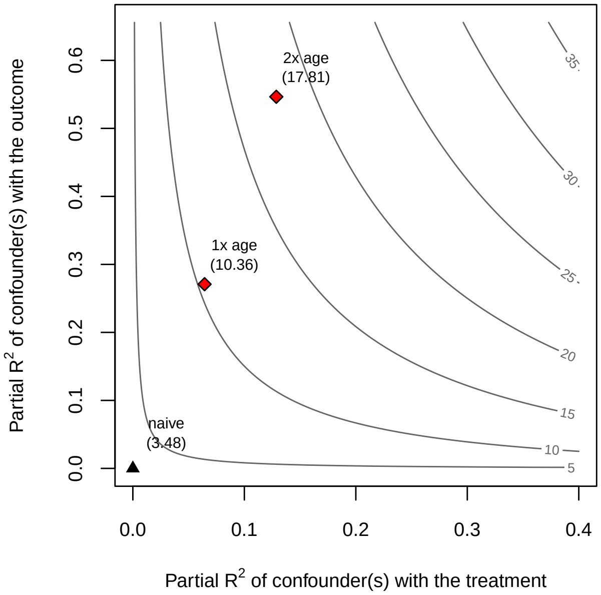

**(a)** *sensemakr* for point estimate

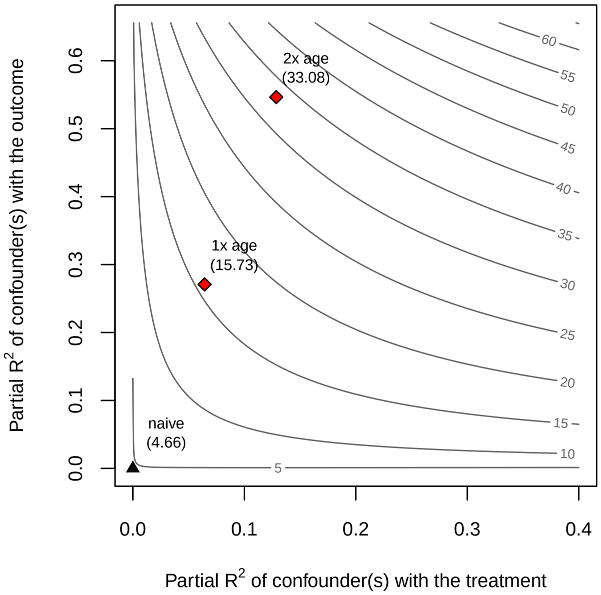

**(b)** *sensemakr* for t-value

**Table S4.** Benchmark upper bounds for sensemakr's bias parameters, $R^2_{X\sim U|C}$ and $R^2_{Y\sim U|X,C}$, based on partial $R^2$ of a measured covariate (or group of covariates) with exposure $X$ and outcome $Y$. Data from the National Health and Nutrition Examination Survey.

| Measured covariate | $R^2_{X\sim U\|C}(\%)$ | $R^2_{Y\sim U\|X,C}(\%)$ |
|---|---|---|
| Age | 6.43 | 27.08 |
| Sex | 0.07 | 1.09 |
| Age and sex | 6.53 | 28.49 |

**Figure S8.** E-value quantitative bias analysis plot for the effect of diabetes on systolic blood pressure from the National Health and Nutrition Examination Survey study. Curves denote bias parameter values that explain away $100\%$ of the naive point estimate (red) and statistical significance at the $1\%$ level (black); dots denote corresponding E-value.

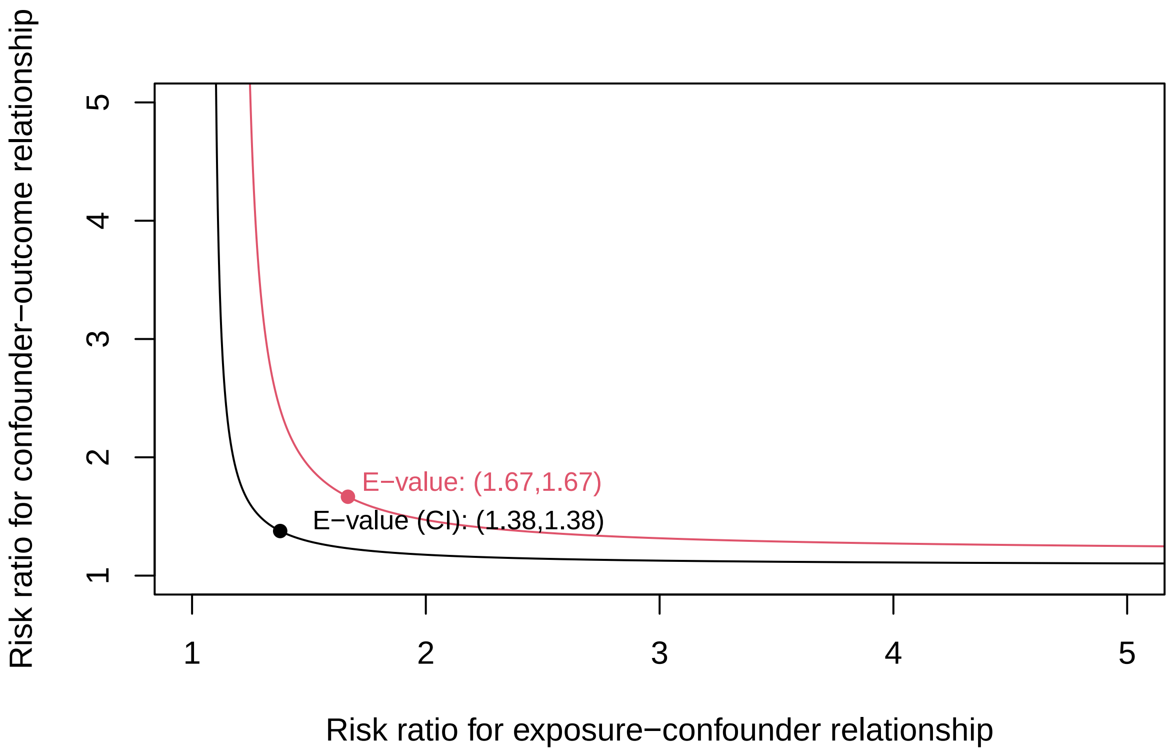

**Table S5.** Benchmark E-values, for the point estimate and lower $99\%$ confidence interval (CI) limit, based on omitting each measured covariate while controlling for the remaining measured covariates. For the effect of diabetes on systolic blood pressure from the National Health and Nutrition Examination Survey.

| | E-value | |
|---|---|---|
| Measured covariate | Point estimate | Lower CI limit |
| Age | 2.61 | 2.27 |
| Sex | 1.69 | 1.40 |

**Figure S9.** *konfound*: Threshold plot representing the percent bias necessary to invalidate inference (at $1\%$ statistical significance) for the effect of diabetes on systolic blood pressure from the National Health and Nutrition Examination Survey study.

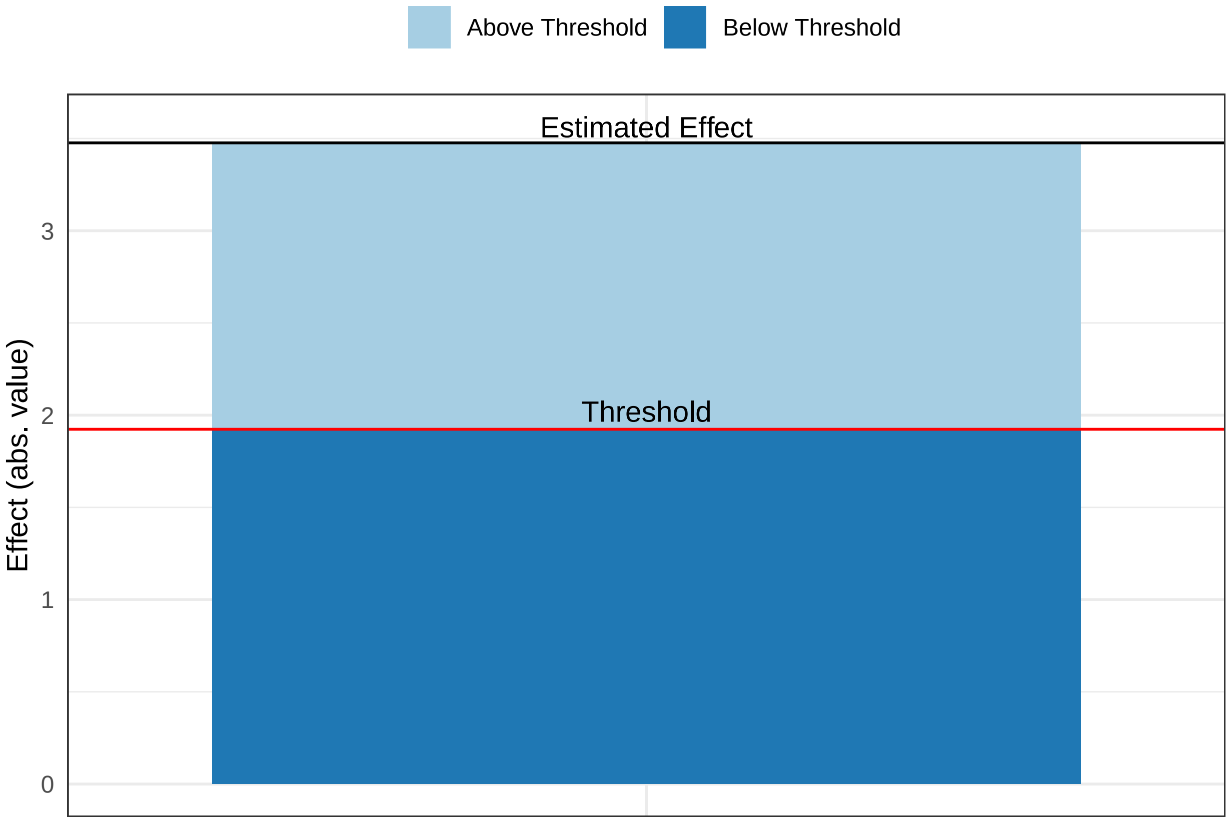

**Figure S10.** *konfound*: Correlation plot of the impact of unmeasured confounding necessary to invalidate inference (at $1\%$ statistical significance) for the effect of diabetes on systolic blood pressure from the National Health and Nutrition Examination Survey study. Partial correlations $R_{x \cdot cv|Z}$ and $R_{y \cdot cv|Z}$ are between unmeasured confounder $cv$ and exposure $X$, and outcome $Y$, respectively, after accounting for measured covariates $Z$.

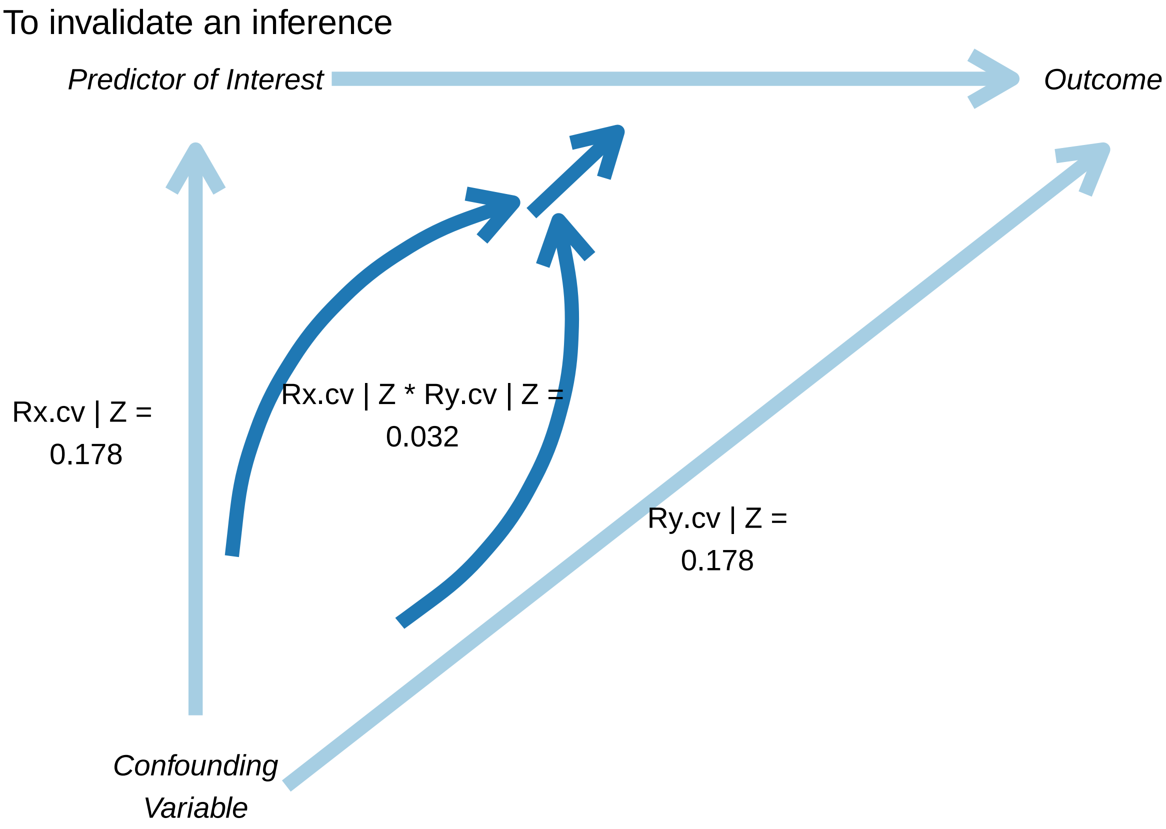

**Table S6.** *konfound*: Benchmark values for $r_{X \sim U|C}$ and $r_{Y \sim U|C}$, and impact threshold $r_{X \sim U|C} \times r_{Y \sim U|C}$, based on partial correlations of each measured covariate with diabetes and systolic blood pressure (SBP). Data from the National Health and Nutrition Examination Survey study.

| Covariate | Partial correlation with diabetes | Partial correlation with SBP | Impact Threshold |
|---|---|---|---|
| Age | 0.25 | 0.46 | 0.12 |
| Sex | -0.03 | -0.11 | -0.33 |

#### The panel study

We conducted a small panel study to obtain feedback on how easy it is to interpret the graphical and tabular output of the five QBA programs: *treatSens*, *causalsens*, *sensemakr*, *EValue* and *konfond*. We invited researchers from the Leiden University Medical Center to participate and provided each participant with a document that contained the QBA output from our analysis of the BCG study. We also asked the participants their experience of a QBA analysis and their prior knowledge of the above five QBA methods.

Seven early career researchers, with up to five years of research experience, agreed to participate. Only one researcher had previously conducted a QBA as part of their own research. With regards to the five QBA methods, two participants had previously heard or read about *causalsens* and *sensemakr* and five had previously heard or read about *EValue*. Not one participant had come across *treatSens* or *konfound*. The researchers were asked to interpret the outputs and comment on the study conclusions. In the absence of benchmark values, participants found it difficult to interpret the output of *EValue*. When referring to the benchmark values to gauge the plausible effect of unmeasured confounding, all participants reached the same conclusions (i.e., only the results from *causalsens* indicated sensitivity of the study conclusions to unmeasured confounding by childhood socioeconomic position). With regards to ease of interpretation, three participants reported difficulties interpreting the output of *sensemakr* and *konfound*, and one participant did not understand the output from program *EValue*. Furthermore, one participant commented that the output from *causalsens* was easier to interpret than that of *treatSens* and another participant reported the reverse.
